# A platform for brain network sensing and stimulation with quantitative behavioral tracking: Application to limbic circuit epilepsy

**DOI:** 10.1101/2024.02.09.24302358

**Authors:** Vaclav Kremen, Vladimir Sladky, Filip Mivalt, Nicholas M. Gregg, Irena Balzekas, Victoria Marks, Benjamin H. Brinkmann, Brian Nils Lundstrom, Jie Cui, Erik K. St Louis, Paul Croarkin, Eva C Alden, Julie Fields, Karla Crockett, Jindrich Adolf, Jordan Bilderbeek, Dora Hermes, Steven Messina, Kai J. Miller, Jamie Van Gompel, Timothy Denison, Gregory A. Worrell

**Author notes:** V.K., F.M, and V.S. contributed equally to this work. Unrelated to this research, G.A.W., B.H.B., J.V.G., and B.N.L are named inventors for intellectual property developed at Mayo Clinic and licensed to Cadence Neuroscience. G.A.W. has licensed intellectual property developed at Mayo Clinic to NeuroOne. Mayo Clinic has received research support and consulting fees on behalf of G.A.W., B.N.L., and B.H.B. from Cadence Neuroscience, UNEEG Medical, NeuroOne, Epiminder, Medtronic, and Philips Neuro. P.C. has received research grant support from Neuronetics, NeoSync, and Pfizer; grant in kind (equipment support) from Assurex, MagVenture, and Neuronetics; and served as a consultant for Engrail Therapeutics, Myriad Neuroscience, Procter & Gamble, and Sunovion. T.D. is a consultant for Synchron, is on the advisory board of Cortec Neuro, is a shareholder collaborator of Bioinduction, and a shareholder director of Amber Therapeutics. All the other authors declare no competing financial interests.

## Abstract

Temporal lobe epilepsy is a common neurological disease characterized by recurrent seizures. These seizures often originate from limbic networks and people also experience chronic comorbidities related to memory, mood, and sleep (MMS). Deep brain stimulation targeting the anterior nucleus of the thalamus (ANT-DBS) is a proven therapy, but the optimal stimulation parameters remain unclear. We developed a neurotechnology platform for tracking seizures and MMS to enable data streaming between an investigational brain sensing-stimulation implant, mobile devices, and a cloud environment. Artificial Intelligence algorithms provided accurate catalogs of seizures, interictal epileptiform spikes, and wake-sleep brain states. Remotely administered memory and mood assessments were used to densely sample cognitive and behavioral response during ANT-DBS. We evaluated the efficacy of low-frequency versus high-frequency ANT-DBS. They both reduced seizures, but low-frequency ANT-DBS showed greater reductions and better sleep and memory. These results highlight the potential of synchronized brain sensing and behavioral tracking for optimizing neuromodulation therapy.

## Introduction

Epilepsy is a common neurologic disease characterized by recurrent seizures and affects over 50 million people worldwide^1,2^. In addition to seizures, the quality of life for people with epilepsy (PWE) can be markedly affected by psychiatric and neurologic comorbidities^3^. Epilepsy is a network circuit disorder with dysregulation of specific brain networks underlying the sporadic seizures and chronic comorbidites^3–5^. Anti-seizure medications are the mainstay of epilepsy therapy, but over 1/3 of PWE are drug resistant^6^ and continue to have seizures despite taking daily medications.

Mesial temporal lobe epilepsy (mTLE) is one of the most common epilepsies and is defined by focal seizures originating from limbic circuits^7,8^ involving amygdala, hippocampus (HPC) and parahippocampal neocortex. It is frequently drug resistant and mood, memory, and sleep (MMS) disturbances^9,10^ are common given the limbic circuit origin. Resective or ablative surgical procedures targeting mesial temporal structures are proven treatments for drug resistant mTLE^10,11^. Many people with mTLE, however, are not good surgical candidates because destruction of the circuits responsible for their seizures will negatively impact memory^10,12–14^. Since the same limbic circuitry responsible for mTLE continues to perform vital cognitive function and emotional regulation when it is not producing sporadic, disabling seizures. People with normal structural imaging^12^, bilateral mTLE^15^, or high baseline memory performance^13^ pose a particular challenge for safe and effective surgery, inspiring interest and need for non-destructive, reversible therapeutic approaches such as electrical brain stimulation.^16^

Deep brain electrical stimulation targeting the anterior nucleus of the thalamus (ANT-DBS) is a proven neuromodulation therapy for focal epilepsy^17,18^. The pivotal SANTE trial was a landmark multi-center, placebo-controlled, double blinded clinical trial that showed duty cycle, high-frequency (145 Hz; 1 minute on and 5 minutes off) ANT-DBS reduced patient reported seizures. Most patients (66%) in the SANTE trial had TLE. The use of high-frequency (HF-ANT) stimulation was motivated by animal research showing it increased seizure threshold^19,20,21^ and disrupted seizure propagation^16,22^

High-frequency ANT-DBS produced a 40.4% median seizure reduction from baseline compared to 14.5% in the control non-stimulation group (p < 0.05) over 3 months. In the subsequent open-label phase the median seizure frequency reduction reached 69%, and 16% of participants had at least one 6-month seizure-free period over the 5-year open label phase^18^. Given the ANT^23,24^ is an important limbic network node it is not surprising that ANT-DBS can impact mood, memory, and sleep (MMS) comorbidities. In the SANTE trial, participants receiving HF-ANT reported depressive mood (14.8% vs. 1.8%) and memory impairment (13.0% vs. 1.8%) symptoms more frequently than the non-stimulated control group^17^. However, there was no significant change at a group level in the standard neuropsychological assessments ^17,25^. The effect of ANT-DBS on sleep has received less attention, but it is also known HF-ANT can disrupt sleep^26–28^.

These results highlight the importance of quantitatively tracking seizures and MMS comorbidities during ANT-DBS. Relatively little is known about ANT-DBS parameter optimization ^29,30^. Patient reported seizure diaries are known to be inaccurate^31–34^, and seizure detection is very limited on currently available brain sensing devices^35–37^. Without accurate seizure catalogs and appropriately dense behavioral tracking of MMS comorbidities, optimizing neuromodulation therapy will remain challenging.

Inaccurate seizure diaries and inadequate sampling of common MMS comorbidities remain fundamental gaps in clinical epileptology. Informed by our previous studies in humans and pet canines with epilepsy^34,38–40^ using streaming hippocampal local field potentials (LFPs) for accurate seizure and interictal epileptiform spike (IES) detection, and motivated to collect continuous ecologically realistic behavioral data we developed a neurotechnology platform enabling the creation of accurate catalogs of seizures, IES, and MMS comorbidities. The BrainRISE (Restoration and Intelligent Stimulation Ecosystem) platform enables bi-directional communication between an implanted sensing-stimulation device, local mobile devices, and cloud computing environment using wireless cellular and internet protocols. Remote assessments of mood, memory and sleep are synchronized with limbic network LFPs. Narrow Artificial Intelligence (AI) algorithms create accurate catalogs of seizures, IES, and wake-sleep brain states.

High-frequency (HF-DBS) was proven effective in the SANTE trial and low-frequency (LF-DBS) has been shown to suppress seizure thresholds in rodent^41–45^ models and in humans^46,47^. To directly evaluate LF-DBS versus HF-DBS on seizures and MMS comorbidities in mTLE we used the BrainRISE platform and a novel investigational sensing-stimulation implantable device with embedded analytics and four electrode leads targeting bilateral amygdala-hippocampus and ANT.

## Results

### Participants and Protocol

Seven people with drug-resistant mesial temporal lobe epilepsy were enrolled under FDA IDE: G180224 *Human Safety and Feasibility Study of Neurophysiologically Based Brain State Tracking and Modulation in Focal Epilepsy* https://clinicaltrials.gov/ct2/show/NCT03946618. All seven participants had co-morbid depression, anxiety, and sleep disturbances (insomnia, hypersomnolence). Participants had comprehensive Phase-I non-invasive evaluations for their drug resistant epilepsy including MRI, DTI, functional imaging, and multi-day video scalp EEG to record their habitual seizures. Three participants had invasive intracranial stereo-EEG. Six participants met the inclusion criteria with at least 3 reported disabling seizures per month at baseline using the mobile application to create an electronic diary of reported seizures. One participant declined further participation after the baseline data collection. Five participants were implanted with an investigational neural sensing-stimulation device (Medtronic Summit RC+S^TM^ system, Figure 1, and supplementary material).

**Figure 1.**
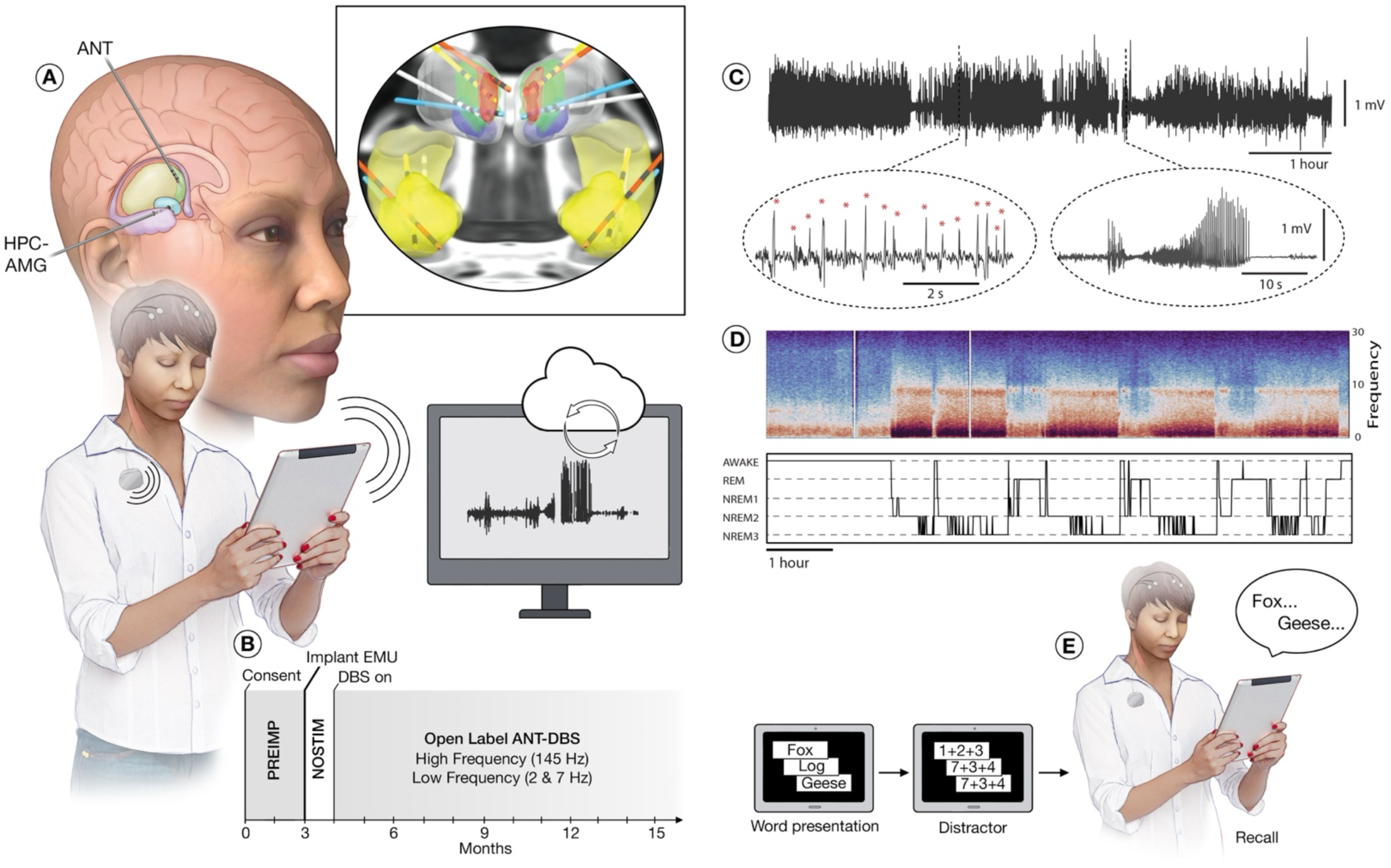
Monitoring brain electrophysiology and behavior in ambulatory humans living in their home environment. **A)** The neural stimulation and sensing implant device enables wireless streaming of brain local field potentials (LFP) to a mobile device running the epilepsy patient assistant application (EPA). The EPA application orchestrates integration and synchronization of multiple wireless devices and provides a system for bidirectional communication between devices, participants, and the clinical team. In participants with mesial temporal lobe epilepsy (mTLE), limbic circuit network nodes were implanted with 4-contact leads targeting bilateral amygdala-hippocampus (HPC) and anterior nucleus of thalamus (ANT) (inset view). The sensing-stimulation lead extensions were tunneled under the skin to the RC+S^TM^ device implanted in a surgically created pocket over the left pectoralis muscle. The RC+S^TM^ device supports bidirectional wireless communications and streaming of 4-bipolar LFP channels to a mobile device via a relay telemetry device typically worn by the individual. Bidirectional wireless communication between the implantable neural sensing-stimulation device, mobile device, and cloud environment enables analysis of synchronized brain electrophysiology, patient inputs, and behavior. Automatically or manually triggered behavioral surveys run on the mobile device. **B)** Safety and feasibility protocol for the 4-lead limbic network implant with investigational neural sensing-stimulation Medtronic Summit RC+S^TM^. **C)** Representative LFP recorded from epileptogenic HPC. Automated narrow Artificial Intelligence algorithms were deployed to capture pathological interictal epileptiform spikes (IES) and electrographic seizure discharges (Sz). Circled insets highlight automated IES and Sz detections in an ambulatory participant (M1) in their natural home environment. **D)** *Top:* Representative LFP spectral power recorded from HPC. The characteristic spectral changes associated with awake and sleep behavioral states are visually evident with increased Delta frequency (1 – 4 Hz) activity in sleep and increased Beta frequency (13 – 20 Hz) activity in wakefulness. Simultaneous scalp polysomnography and LFP recordings were used to develop an automated LFP based behavioral state classifier: awake (W), rapid eye movement (REM) sleep, and non-REM sleep. *Bottom:* Hypnogram generated from continuous LFP recordings using a Naïve Bayesian Classifier. **E)** Verbal memory tasks and mood assessments were performed remotely in participants’ home environment. Abbreviations: non-rapid eye movement sleep (NREM), rapid eye movement sleep (REM), investigational Medtronic Summit RC+S^TM^ (RC+S^TM^).

The primary study outcome measures were: **1)** Device-related adverse events (AE) with investigational 4-lead Medtronic Summit RC+S^TM^ system (RC+S^TM^) integrated with off-the-body local and cloud computing. The goal was to show device-related AE with a 4-lead sensing and stimulation device with electrode leads implanted in bilateral amygdala-hippocampus and ANT is not greater than observed in the pivotal SANTE trial that used only 2-leads targeting bilateral ANT^17^. **2)** Feasibility of 24/7 continuous LFP streaming. **3)** Feasibility of creating accurate catalogs of seizures, IES, mood, memory, and wake-sleep behavioral state.

The creation of precise, comprehensive catalogs of seizures, IES, wake-sleep behavior, and neuropsychological mood and memory data was achieved by integrating multiple devices using wireless data streaming protocols with cloud computing infrastructure and artificial intelligence (AI). We used this system to investigate the effect of low-frequency (LF-DBS) and high-frequency (HF-DBS) paradigms on IES, seizures, sleep, mood, and memory.

### BrainRISE Platform Performance

The BrainRISE (Restoration and Intelligent Stimulation Ecosystem) platform enables bi-directional communication between an implanted sensing-stimulation device, mobile devices, and cloud computing environment using wireless cellular and internet protocols^38^. The connectivity between devices is orchestrated with a custom software application (Epilepsy Patient Assistant: EPA) operating on a mobile computing device carried by the participant^48^. The system uses U-band and Bluetooth wireless protocols to maintain connectivity between the multiple wireless capable devices and cloud computing environment (see supplementary material). The platform enables connection of wearable devices to collect rich behavioral data and orchestrate scheduled and responsive remote questionnaires launched based on LFP defined event detections.

Continuous HPC sensing and LFP streaming provides the input signal for validated automated narrow AI algorithms to create accurate catalogs of seizures, IES^34,49^ and sleep-wake behavioral state^39,40,50^ (Figure 1 & 2, and see supplementary material).

Participants kept three devices charged to wirelessly stream LFP data: 1) RC+S^TM^ neural sensing and electrical stimulation implanted devices; 2) telemetry antennae; 3) mobile device. The ability to manage the 3 devices varied across participants. On average, we recorded 99.2 ± 43.0 weeks of data from each patient during ANT-DBS. The average performance for bidirectional streaming data flow was as high as 13.84 hrs. per day.

### Outcomes & System Related Adverse Events

Five people with mTLE were implanted with bilateral ANT leads and bilateral amygdala-hippocampal (AMG-HPC) leads. The ANT leads were used for ANT-DBS and the AMG-HPC leads were used for LFP sensing. Each implanted lead has 4 electrode contacts that can be used for sensing or electrical stimulation (Figure 1 and supplementary materials). All five participants had independent left and right temporal lobe IES and seizures captured during the phase-I non-invasive evaluation. Participants M1, M3, and M4 had phase-II invasive evaluations with intracranial stereo-EEG demonstrating bilateral independent IES and seizures.

The custom epilepsy patient assistant (EPA) application running on a mobile device was used by participants to report their seizures. When compared to baseline reported seizure counts the patient-reported seizures were reduced with both HF-DBS and LF-DBS stimulation (Figure 2A; p < 0.05). The magnitude of reported seizure reduction from baseline using LF-DBS and HF-DBS paradigms was similar (−52% and −56%, p > 0.05).

**Figure 2:**
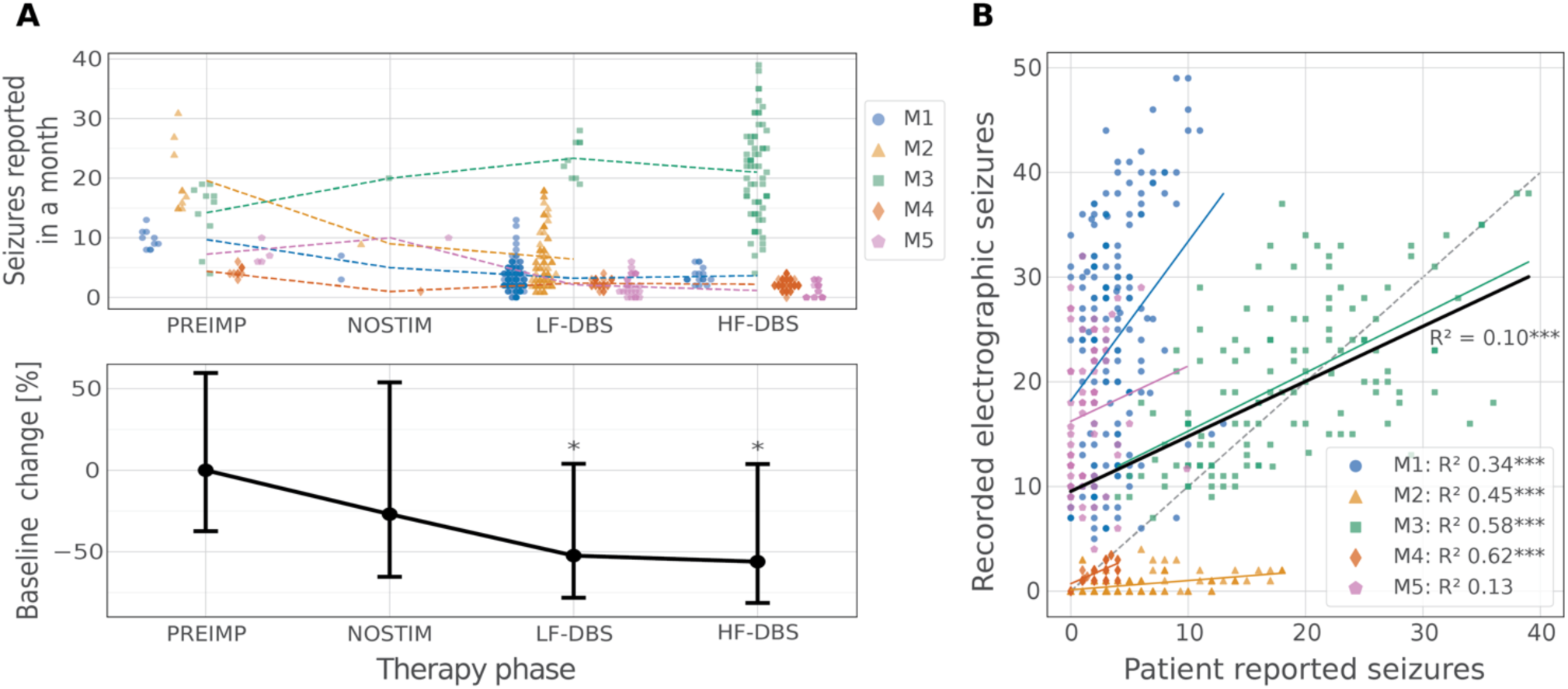
Anterior Nucleus of Thalamus Deep Brain Stimulation (ANT-DBS) and Seizures. Patients reported their seizures using the epilepsy patient application (EPA) running on a mobile device during baseline prior to surgery (PREIMP: 3 months prior to implant), after device implant before starting ANT-DBS (NOSTIM: 4 weeks), and during low-frequency (LF-DBS) and high-frequency (HF-DBS). **A)** *Top:* Scatter plot of individual monthly reported seizures and the mean over all months (dashed lines). *Bottom:* Both LF-DBS and HF-DBS reduced reported seizures compared to baseline by over 50% at a group level within a General linear mixed model (GLMM, p < 0.05), and in four of five individuals. The patient (M4) with an increase in reported seizures after implant was receiving high-dose diazepam therapy that had to be discontinued because of mood decline and fatigue on week 5 after implant, which may have precipitated an increase in seizures. **B)** Patient reported seizures versus detected electrographic seizures captured with continuous hippocampal local field potentials (LFP) streaming. Notably, some participants reported seizures (e.g. M3) that were not associated with an electrographic LFP correlate and some (M1&M5) had more electrographic LFP seizures than they reported. *(See supplementary materials)*

In the five mTLE participants implanted with the RC+S^TM^ device comprehensive mood, quality of life, seizure severity, and neuropsychological testing during in person clinic visits were collected at pre-implant baseline, 3 - 6 months post-implant, and at 9 - 15 months post implant. There were no significant differences across participants and timepoints for the standard measures of mood, quality of life, seizure severity, or neuropsychological (NP) testing (supplementary materials).

Four device related adverse events were reported over the 15-month protocol. Participant M2 inadvertently disabled therapy using the EPA application. A subsequent EPA reversion corrected this potential for operator mistake, and the cloud dashboard more clearly displayed the therapy status to the clinical team. Two participants (M1 & M4) experienced behavioral changes with anxiety, dysphoria, and sleep disturbance during trials of continuous HF-DBS (see supplementary data). In both participants the reported anxiety, dysphoria and sleep disturbance resolved with changing from continuous HF-DBS to duty cycle (1 min on & 5 min off; SANTE parameters^17^). In participant M5 the right amygdala-hippocampal lead could not be fully seated into the lead extension connector at the time of surgery, leaving only three of the four contacts recording. There were no deaths, infections, bleeding, strokes, status epilepticus or other device related complications. All subjects had NP testing at baseline, during LF-DBS and HF-DBS. Participant M3 had a decrease in VPA immediate and delayed recall compared to baseline. For M5 both Rey Auditory Verbal Learning Test (RAVLT) and Verbal Paired Associates (VPA) immediate recall was reduced during high frequency ANT-DBS. These changes did not reach significance.

Subject M4 was hospitalized for whole-body paralysis and unresponsiveness lasting hours that did not have any LFP correlate and was determined to be consistent with a new onset functional nonepileptic spell. Subject M5 had three overnight hospitalizations with increase in recurrent seizures like his baseline.

Participants captured an average of 162 *±* 158 seizures over the course of the study, with variable discrepancies between patient reported seizures and reports with LFP correlates. We directly compared reported seizures with detected electrographic seizures captured with automated AI applied to the continuous LFP recordings^34^ (Figure 2B). There was a correlation between reported seizures and electrographic seizures on a group level, with substantial variability across participants (overall R^2^ = 0.1, p < 0.001). However, participants variably reported seizures without LFP correlates, and under reported detected electrographic seizures (Figure 2B). Participant M4 had the highest correlation between electrographic LFP detected seizures and reporting (R^2^ = 0.62, p<0.001). This is in contrast with the absence of correlation in participant M5 (R^2^ = 0.13, p>0.05). It is noteworthy that although participant M4 was amnesic for her focal impaired awareness seizures (FIAS), her spouse was often able to accurately log these episodes. This was made possible due to the increased and closer observation during the work-from-home arrangements during the COVID-19 lockdown.

### Seizures and Interictal Epileptiform Spikes and their Circadian Patterns

All patients had seizures start unilaterally in AMG – HPC, and then propagate to contralateral AMG - HPC. All participants had bilateral independent IES, and all but M4 had bilateral independent seizures recorded. In all participants the IES activity was increased in left compared to right AMG-HPC. Similarly, M1, M2, M4 and M5 had more seizures originating from the left AMG-HPC (supplementary materials).

Characterization of long-term LFP recordings showed that electrographic seizures and IES occurred with greater frequency in the left HPC in each of the 5 participants. All participants had day-night patterns of epileptiform activity, with IES maximal at night during sleep and seizures maximal during the day during wakefulness (Figure 3). The seizures captured from continuous LFP recordings demonstrated a strong diurnal pattern with most seizures occurring during the daytime (823 seizures during wakefulness vs. 113 seizures during sleep (p<0.0001), supplementary data). The timing of the daytime seizures was patient specific, with M4 showing a tendency for seizures at 2:30 PM ± 1 hr., two participants (M1, M3) with seizures preferentially occurring in the morning and evening, M2 with late afternoon and evening seizures, and M5 with more uniform distribution of seizures over the day.

**Figure 3.**
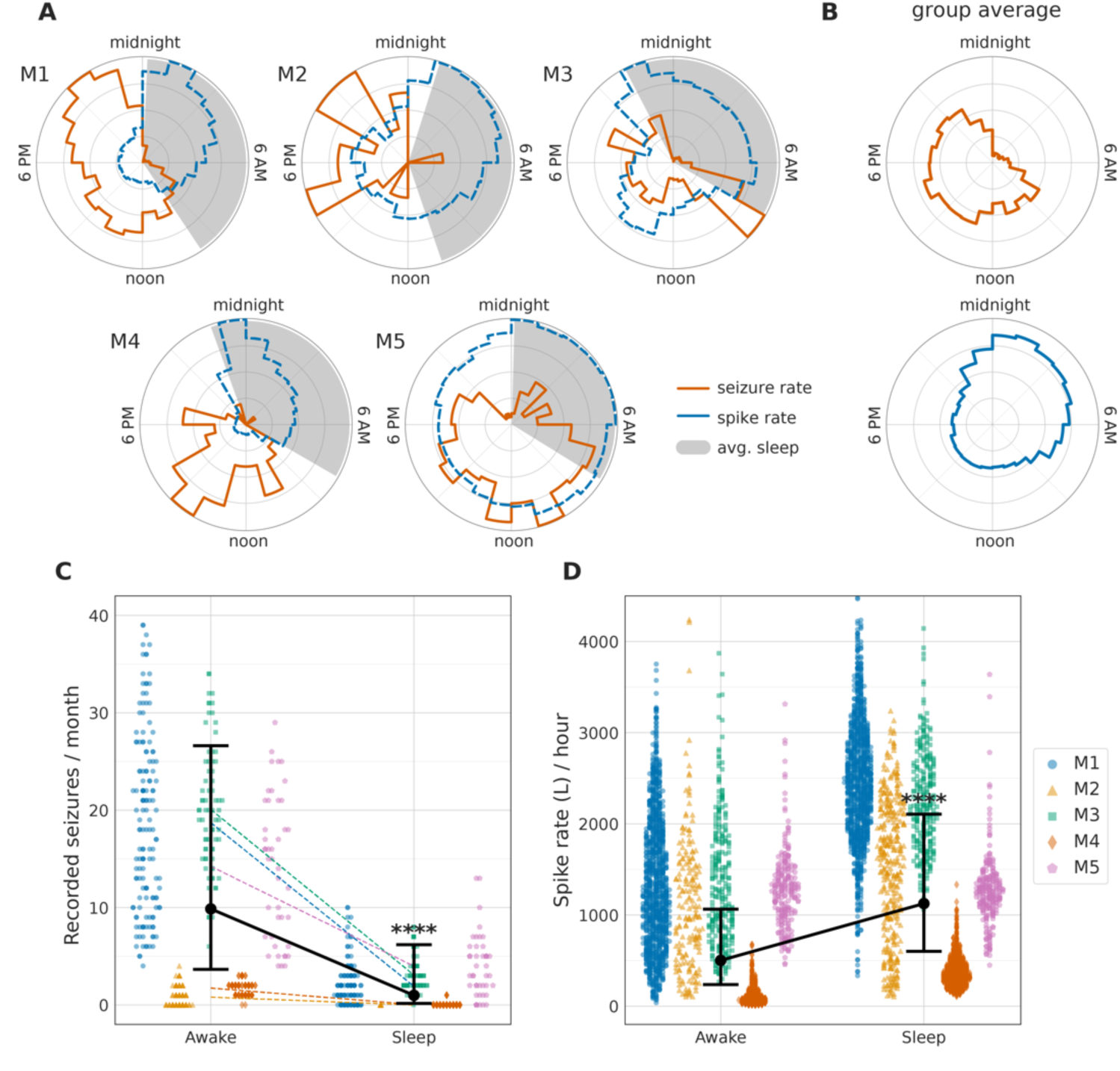
Circadian Patterns of Seizures and Interictal Epileptiform Spikes. Epileptogenic human hippocampus (HPC) generates spontaneous seizures (Sz) and interictal epileptiform spikes (IES) that are modulated by awake-sleep behavioral states. The habitual Sz primarily occurred diurnally during wakefulness with a complex, patient specific, distribution of increased occurrence. Seizures rarely occurred during sleep compared to wakefulness. Conversely, IES were increased during sleep, particularly in non-rapid eye movement (NREM). **A)** Circular 24-hr histograms of Sz occurrence for each participant show detected electrographic Sz (red-solid), IES rates (blue-dashed) and sleep-wake classification (gray-filled) using validated algorithms applied to continuous local field potentials (LFP). Everyone’s Sz primarily occur during wakefulness with patient specific distributions approximating unimodal (M4 & M5) or bimodal (M1,2,3) daytime distributions. **B)** Aggregated Sz and IES across all patients show the differential behavioral state modulation of epileptiform activity, IES and Sz. **C)** The seizures are more common in daytime during wakefulness (p<0.0001) and **D)** IES rates are higher in sleep (p < 0.0001). The General linear mixed model group estimated fit with confidence intervals in solid black.

The pattern of IES over a 24-hr period is distinctly different from seizures. The IES rates are increased during sleep compared to wake for all participants (p<0.0001) when seizures are less likely to occur. (Figure 3B&C and see supplementary material).

### Impact of ANT-DBS on Detected Seizures and IES

The number of reported seizures decreased with ANT-DBS when compared to baseline for both HF-DBS and LF-DBS (Figure 2A). The seizures detected from streaming LFP, however showed that LF-DBS reduced seizures more than HF-DBS (p < 0.05). Furthermore, LF-DBS significantly reduced IES compared to HF-DBS during both sleep and wakefulness (p < 0.01) (Figure 4). Seizures detected from streaming LFP were similar for HF-DBS and LF-DBS during sleep, but the number of seizures during sleep were small (p > 0.05).

**Figure 4:**
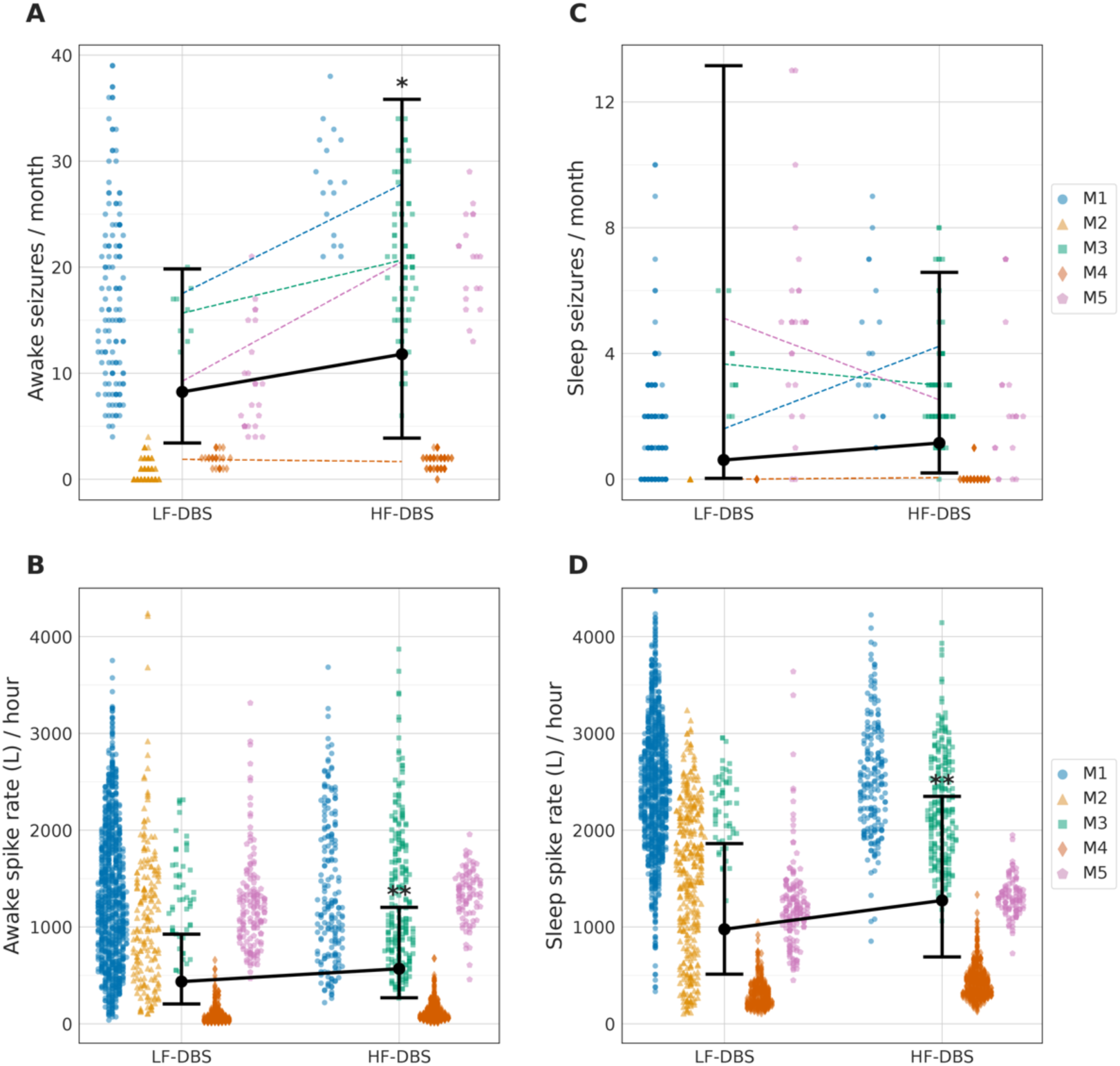
Electrographic seizures and interictal epileptiform spikes (IES) recorded from hippocampus (HPC) during Anterior Nucleus Thalamus Deep Brain Stimulation. A validated automated seizure and IES detector was used to catalog electrographic seizures (Sz) and IES from streaming hippocampal (HPC) local field potential (LFP) recordings (see also Figure 1B) over multiple months in participants with mTLE. Here we compare low frequency (LF-DBS) and high frequency (HF-DBS) stimulation paradigms. The GLMM group estimated fit with confidence intervals in solid black. **A)** The electrographic seizures during wakefulness recorded from bilateral HPC were reduced during continuous LF-DBS compared to duty cycle HF-DBS. **B)** The IES rates during wakefulness were decreased during LF-DBS compared to HF-DBS for all 5 patients. **C)** Electrographic seizures during sleep recorded from bilateral HPC are infrequent and similar during LF-DBS and HF-DBS. **D)** The IES rates during sleep were decreased during LF-DBS compared to HF-DBS (p < 0.01).

### Impact of ANT-DBS on Sleep

The LFP recordings from HPC were used as input to a validated, individualized, automated Naive Bayes sleep-wake state classifier^39^ (see methods and supplementary data). As participants slept ad libitum on their own preferred sleep-wake schedules, characteristics of sleep varied widely with a range of individual features for sleep onset, offset, and duration. The number of NREM-REM cycles demonstrated substantial intra-individual and inter-individual variability. On average, across all participants, the total sleep duration was 8 hrs. 33 minutes, with 4.04 NREM-REM cycles and 19.9 min NREM epoch duration.

ANT-DBS neuromodulation with LF-DBS and HF-DBS did not impact total sleep duration (sleep offset – onset), but the time spent awake after sleep onset (WASO) was reduced by LF-DBS compared to baseline without stimulation (p < 0.05). In contrast, HF-DBS increased WASO compared to baseline and LF-DBS (p < 0.001). The overall time spent in NREM and REM was decreased during HF-DBS compared to baseline without stimulation and LF-DBS. The total NREM-REM time in sleep with LF-DBS neuromodulation was indistinguishable from baseline without DBS and longer than HF-DBS (p < 0.05). (Figure 5 & supplementary materials).

**Figure 5:**
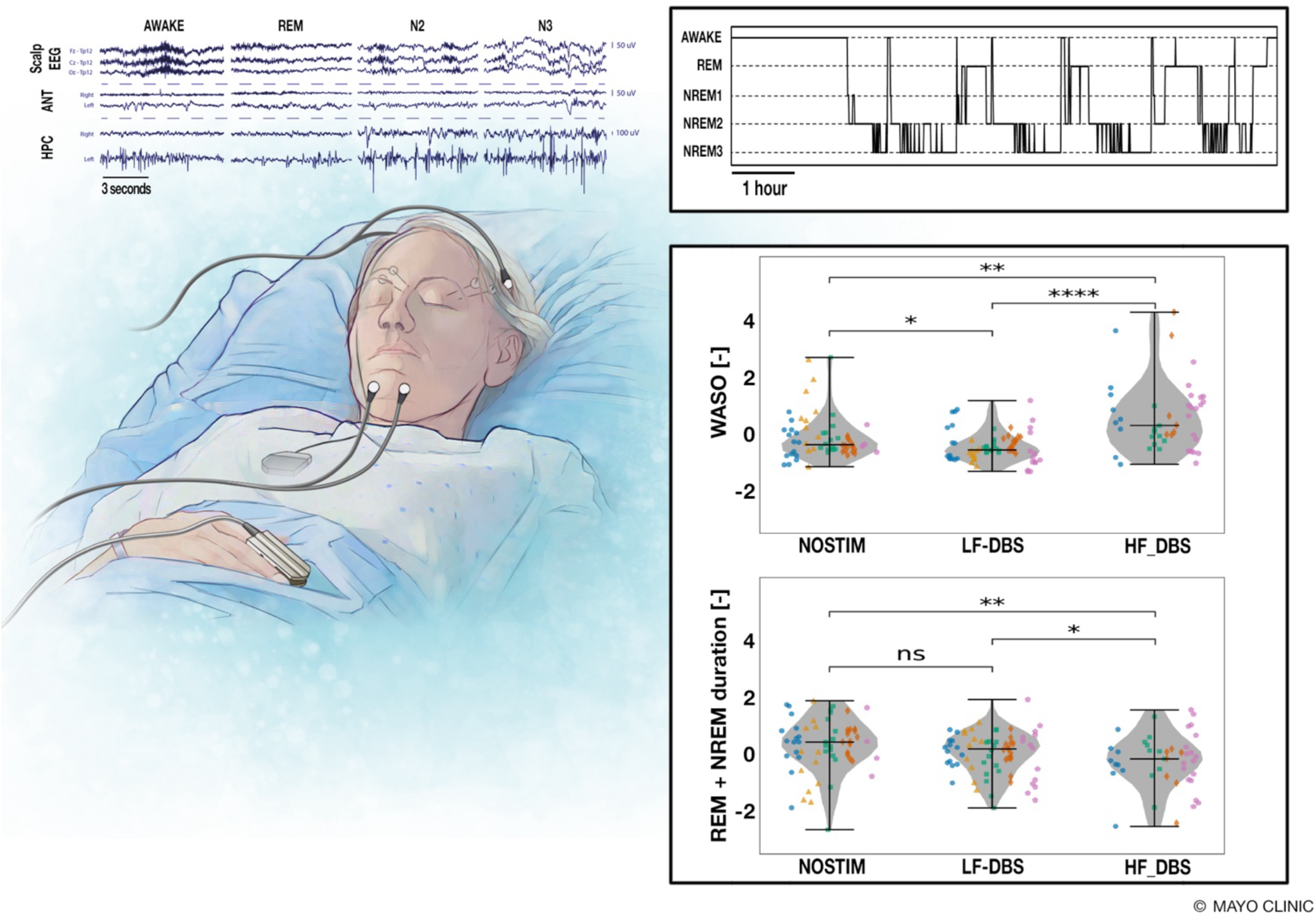
Effect of Anterior Nucleus of Thalamus Deep Brain Stimulation (ANT-DBS) on Sleep. The impact of ANT-DBS neuromodulation on sleep was investigated using long-term behavioral state classifications (awake-sleep) from a validated Naïve Bayesian Classifier applied to local field potentials (LFP) recordings from the hippocampus (HPC) of ambulatory participants in their home environments. **Top Left)** Simultaneous polysomnography and streaming intracranial LFP recordings are used for training and testing of the automated sleepwake classifier. Top Right) An example of a gold-standard hypnogram during simultaneous polysomnography and intracranial LFP recording. Similar hypnograms were obtained for multiple months using only intracranial LFP in participants in their home environments. The automated wake-sleep classifier labeled behavioral states as Wakefulness (W), rapid eye movement sleep (REM) and non-REM sleep (NREM). Bottom Right) The time spent awake after sleep onset (WASO) was reduced by LF-DBS compared to baseline without stimulation (p < 0.05). HF-DBS increased WASO compared to both baseline (p < 0.01) and LF-DBS (p < 0.0001). The overall time spent in NREM and REM was reduced during HF-DBS compared to the no stimulation baseline and LF-DBS. The total NREM-REM time in sleep with LF-DBS was indistinguishable from the no stimulation baseline and longer than HF-DBS (p < 0.05).

### Impact of ANT-DBS on Memory (Ambulatory Free Recall Task)

Participants used the epilepsy patient assistant (EPA) application on a mobile device to remotely perform a validated word-free recall task^51–54^ in their natural home environment (Figure 6A). The task was performed with the subject at different times at the participant’s preference (hour, day of week) over multiple months during the two ANT-DBS stimulation paradigms. The ambulatory free-recall verbal memory task was not functional on the EPA system until after the time of implant and no pre-implant baseline data were captured. However, as noted previously all participants had standard NP testing that did not show a change during baseline, LF-DBS, or HF-DBS. All participants performed the ambulatory free-recall memory task after implant of the DBS system. The free-recall memory scores were better during LF-DBS compared to HF-DBS (p < 0.001; Fig. 6B) (See supplementary materials).

**Figure 6:**
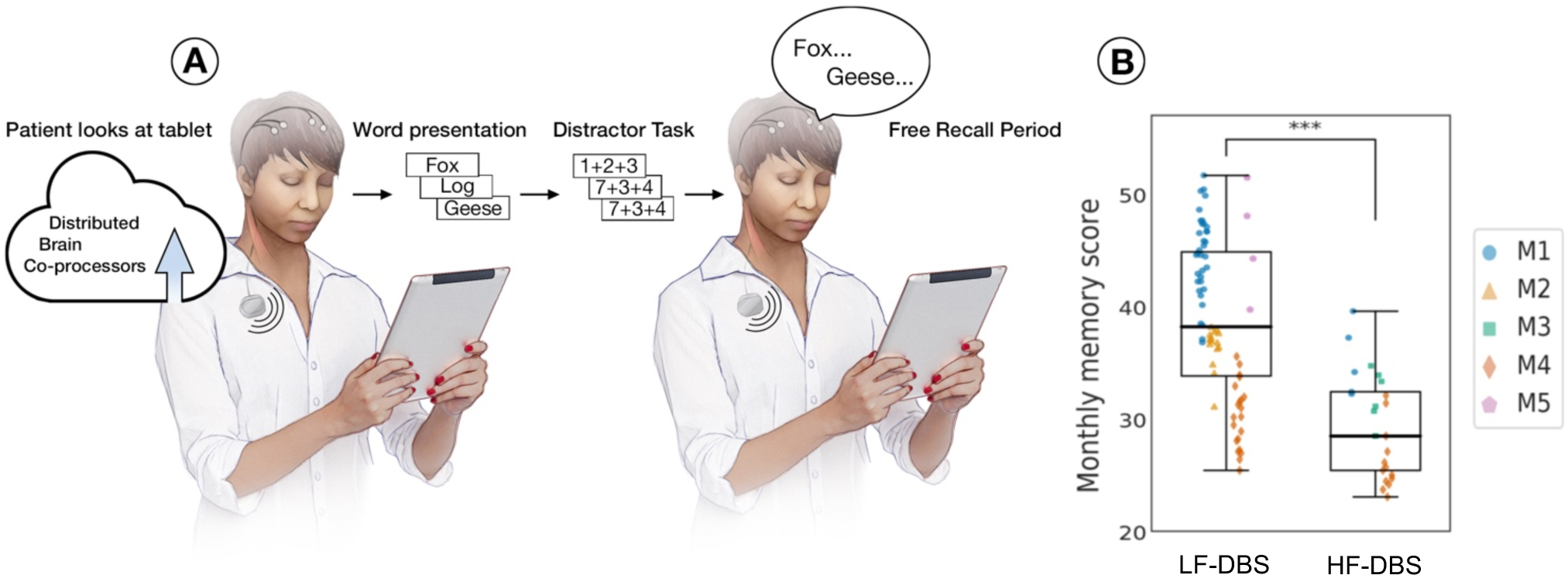
Effect of Anterior nucleus thalamus deep brain stimulation (ANT-DBS) on Verbal Memory Performance. The impact of low-frequency (LF-DBS) and high-frequency (HF-DBS) on verbal memory was investigated using a simple word encoding-recall task performed remotely by participants in their home environment. **A)** Patients used the epilepsy patient assistant (EPA) application running on their mobile device to perform a validated word-recall task. The task was performed with the participant at different times of the day according to their individual preference. The task consisted of an average of 15 trials performed using lists of 12 words. A proper noun is randomly selected from a large database and presented on the screen for 10 seconds. After the presentation of the 12 nouns, a simple math distractor is presented that the patient performs. Subsequently, there is a period of free recall where the patient recalls as many words, in any order, as possible from the prior encoding list. **B)** Verbal memory scores for free recall were better during periods where the participants were receiving LF-DBS compared to HF-DBS. Note, the ANT-DBS is off while the task is being performed.

### Impact of ANT-DBS on Mood

All participants completed assessments of mood (Beck Depression Index) and suicidality (Columbia Suicide) during baseline, LF-DBS and HF-DBS. In addition, during LF-DBS and HF-DBS participants were prompted at random times of the day to complete the Immediate Mood Scaler 12 (IMS)^55^ deployed on the EPA application to densely track mood and anxiety symptoms instantaneously in the naturalistic environment (see supplementary materials). Symptoms of depression and anxiety were present in all five participants, but there were no differences detected between HF-DBS and LF-DBS for mood and anxiety symptoms (see supplementary materials).

## Discussion

Epilepsy is a complex and heterogenous disorder with multiple etiologies. Here we investigated five people with drug resistant mTLE to focus attention on limbic network circuits generating disabling seizures and MMS comorbidities. We created a novel platform (BrainRISE) integrating implantable and wearable devices, and cloud infrastructure to investigate the impact of ANT-DBS therapy on seizures, IES, and MMS. Synchronized data streams^56^ from multiple devices (brain sensing-stimulation implanted device, mobile computing and wearable devices) was streamed to a cloud-based data storage, viewing, and computing environment. Validated narrow AI algorithms were applied to streaming hippocampal LFP recordings to create accurate catalogs of seizures, IES, and wake-sleep behavioral states. Subjects reported seizures, and remotely performed a free recall memory task and answered mood questionnaires using the custom mobile application.

The study demonstrates the feasibility and potential clinical utility of continuous, synchronized tracking of brain electrophysiology and behavior in people with epilepsy living in their natural home environments (Figure 1). We characterized the circadian patterns of epileptiform activity in mTLE and show that seizures occur primarily during wakefulness and IES are markedly increased during sleep. The mechanism underlying the discordance of behavioral state dependent IES and seizures, two fundamental measures of pathological brain excitability, remains unclear. We speculate that thalamo-hippocampal circuits and state-dependent large scale and local circuit neuronal ensembles interact to modulate the local activation, spread and network propagation of pathological hippocampal electrographic activity, with the net effects rendering seizures less frequent during NREM and REM sleep compared to wakefulness.

Indeed, we can quantify the effects of ANT-DBS on mTLE and surprisingly show that LF-DBS is as effective as the FDA approved HF-DBS for reducing patient reported seizures. But importantly, LF-DBS showed greater reduction of electrographic seizures and IES, objective biomarkers of pathologic network excitability^57,58^, in both wakefulness and sleep, and resulted in better objective measures of sleep and verbal memory compared to HF-DBS (Figures 2, 4–6).

HF-DBS is proven to reduce seizures^17^, but there may be more optimal stimulation paradigms depending on the circuits and etiologies underlying the epilepsy, seizures, and MMS comorbidities. Here we show that in a small number of well characterized mTLE participants having hippocampal onset seizures, LF-DBS was more effective in reducing epileptiform activity and was better for sleep and memory than HF-DBS. The clinical relevance of reducing IES in epilepsy has long been debated, but recent studies showed a deleterious impact of IES on memory^59–62^ and sleep^63^. Furthermore, the role of IES on progression and maintenance of epilepsy remains poorly understood, but IES in slow-wave sleep may play a role in consolidation of epilepsy engrams and epileptogensis^64,65^. It is important to note that participants receiving ANT-DBS, both LF-DBS and HF-DBS, over multiple months did not show evidence for kindling with increasing IES and seizures^66^.

The BrainRISE platform used in this study should prove useful for future brain stimulation optimization investigations, targeting not only seizures but IES and related epilepsy MMS comorbidities. Future studies investigating adaptive and responsive electrical stimulation paradigms targeting multiple limbic network nodes in both thalamus and hippocampus are now feasible. The potential for direct translation of a more optimal paradigm to the thousands of PWE implanted with ANT-DBS is very attractive but will require replication and validation in a larger cohort of patients.

There are several limitations in this study. 1) The trial design is focused on safety and feasibility and does not include blinding or randomization of control no stimulation and ANT-DBS therapy. Given the proven, class-I evidence for ANT-DBS we did not feel there was clinical equipoise to withhold ANT-DBS therapy from patients for an extended period. We did not control for duration of the therapy periods, but comparing equal reporting periods did not affect the results. 2) The existence of cycles of seizure occurrence in epilepsy^67^ is now well established, and the results obtained in any trial will likely depend on the timing of therapy and data collection. We believe we have addressed this limitation here with the dense long-term monitoring. 3) Similar to other studies we use patient reports of seizures to assess the outcome of ANT-DBS. There is strong evidence that many PWE are poor reporters of their seizures. This was demonstrated in our data when comparing proven electrographic discharges with patient reports. This is a fundamental limitation in clinical epileptology, but here this gap is partially mitigated by the unique long-term continuous LFP sensing and automated seizure detection after device implant. 4) We obtained standard neuropsychological and memory testing during in person clinic visits at baseline prior to implant and during both LF-DBS and HF-DBS that did not show changes. But we were not able to obtain baseline ambulatory free-recall memory tasks and IMS assessments prior to implant because the software system was not completed prior to enrollment of participants. Therefore, we could only compare differences in memory free recall tasks and IMS scores after implantation between LF-DBS and HF-DBS. 5) The BrainRISE platform provides synchronized behavior and neural activity on an unprecedented scale in ambulatory humans, but challenges remain given the participant burden associated with keeping all device component batteries charged and wireless connectivity active. 5) It remains unknown if, and to what degree, electrographic seizures detected from streaming LFP impair consciousness. This is a fundamental gap in epileptology will be addressed in the future with assessments that are triggered on seizure detections in real time.^68–70^

## Methods

People with drug-resistant mesial temporal lobe epilepsy were identified under FDA Investigation device exemption clinic trial: IDE G180224 and Mayo Clinic IRB: 18-005483 *Human Safety and Feasibility Study of Neurophysiologically Based Brain State Tracking and Modulation in Focal Epilepsy* (clinicaltrials.gov NCT03946618). Here we report the study outcome measures: 1) Adverse events experienced during the IDE. 2) Feasibility of continuous tracking of ANT and HPC local field potentials in ambulatory human participants. 3) Continuous tracking and outcomes of seizures, interictal epileptiform spikes (IES), sleep, mood, and memory during HF-DBS and LF-DBS.

### Participants

Seven participants with drug resistant mesial temporal lobe epilepsy provided written consent in accordance with IRB and FDA requirements. Six participants met the full inclusion criteria with adequate numbers of baseline seizures, and five participants were implanted with an investigational neural sensing and stimulation device (investigational Medtronic Summit RC+S^TM^). The demographics and clinical information of the five participants implanted with the RC+S^TM^ are provided in supplementary materials.

### Outcome Assessments

All participants reported their seizures using a custom mobile application that is part of the BrainRISE platform. In addition to seizure assessments, all participants underwent assessments for mood, quality of life, seizure severity, sleep and neuropsychological (NP) testing. Outcome assessments were obtained at baseline, 3-6 months post-implant, and 9-15 months post-implant. The instruments collected include:

1) Change in mood was assessed by Beck Depression Inventory II & Columbia Suicide Severity Rating Scale (C-SSRS)
2) Change in health-related quality of life (QOL) was assessed by Quality of Life in Epilepsy-31 (QOLIE-31) and SF-36 Health Survey (SF-36).
3) Seizure Severity was assessed using the Liverpool Seizure Severity Scale (LSSS), Seizure Severity Questionnaire (SSQ) Version 3 and Mayo Seizure Scale.
4) Neuropsychological Assessments included: 1) Boston Naming Test, 2) Controlled Oral Word Association Test (COWA), 3) Rey Auditory Verbal Learning Test, 4) Wechsler Memory Scale (WMS-IV) Visual Reproduction Test 5) Wechsler Memory Scale (WMS-IV) Verbal Paired Associates

In addition to the above assessments collected during in person clinic visits with the participant we used BrainRISE platform to remotely collect dense synchronized behavioral assessments and brain activity from ambulatory participants living in their natural home environment.

### BrainRISE Platform

The electrophysiology data^34,38^, patient reports, verbal memory testing^51^ and immediate mood scaler (IMS)^55,71^ were collected using epilepsy patient assistant (EPA), a custom software application running on a mobile computer enabling bidirectional communication between implanted devices with wireless connectivity and local and cloud computing resources. The EPA orchestrates communication between multiple wireless capable devices (implant, mobile device, iPhone, Apple Watch) and features custom automated python narrow AI algorithms for continuous analysis of long-term LFP data, control of electrical stimulation, impedance testing, LFP analysis. The EPA running on a mobile device provides an interface for performing tasks, and collecting patient interactions^38,48^ (Figure 1 and supplementary data). A key aspect of this platform is the application to monitoring ambulatory subjects in their natural home environment, thereby capturing more realistic and ecologically relevant data.

The RC+S^TM^ implanted device brain sensing, electrical stimulation and embedded analytics^72^ do not require continuous connectivity with the BrainRISE system for therapy to remain active. The DBS therapy remains active as long as the implanted RC+S^TM^ device battery is adequately charged. During combined DBS and continuous 4-channel LFP streaming (250 Hz sampling) the RC+S^TM^ device requires daily charging. At 30% battery status the LFP streaming is automatically disabled by the EPA application to maintain therapy. In this scenario with LFP streaming disabled, DBS therapy remains active for approximately 1 week. The system also supports duty cycle LFP streaming paradigms, e.g. 10 minutes every 60 minutes, that can be used to preserve battery charge.

### Investigational Medtronic Summit RC+S^TM^

(supplementary data). In this study the investigational Medtronic Summit RC+S^TM^ implantable device was integrated with the BrainRISE platform. The investigational Medtronic Summit RC+S^TM^ provides bidirectional wireless communication, programmable 16 channel electrical stimulation, and continuous 4 channel (selected bipolar pairs) wireless LFP streaming^38,72,73^. Bipolar pairs of contacts in ANT and AMG-HPC were used for sensing and streaming LFP (sampled at a frequency of 250 – 1000 Hz) to the local mobile device running the custom EPA application. The electrode contacts used to create lead specific bipolar recordings were selected by visual review of LFP data during seizures, resting wakefulness, and sleep-wake transitions.

### Lead and Electrode Contact Localization

(Figure 1A and supplementary material). Five participants (M1, 2, 3, 4, 5) had four leads stereotactically implanted into bilateral ANT and bilateral AMG-HPC. Participants M1, 3, 4, and M5 were implanted with Medtronic 3387-lead in the ANT and Medtronic 3391 in the AMG-HPC sites. Participant M2 had a Medtronic 3387-lead implanted in the left HPC because she had previously undergone a left anterior temporal lobectomy leaving only a residual left HPC tail. Participant M5 does not have a right AMG electrode contact because the 3391-lead tail could not be fully seated into the lead extension connector at the time of surgery, leaving only 3 of the 4 contacts available for recording. For all participants 4 implanted leads (16 total electrode contacts) were localized with post-operative CT scan co-registered to the pre-op MRI and DTI for anatomic localization using previously described pipelines^74,75^. The CT scan and electrode contact positions were co-registered to a T_1_ weighted anatomical MRI scan using SPM12 (https://fil.ion.ucl.ac.uk/spm/)^76^ Freesurfer (http://surfer.nmr.mgh.harvard.edu/) was used to segment the T_1_ weighted MRI and the electrodes labeled according to the Destrieux atlas^77,78^. The final electrode contact localization for impedance analysis was performed with Lead DBS^75^.

### Epilepsy patient assistant (EPA)

The electrophysiology data and patient reports were collected using epilepsy patient assistant (EPA), a custom software application running on a mobile device enabling bidirectional communication between the implanted device and local and cloud computing resources. The EPA features include validated automated algorithms for continuous analysis of LFP data, control of electrical stimulation and impedance testing, LFP analysis and an interface for collecting patient interactions.^38,48^

### Electrical Stimulation of ANT

The ANT electrode pairs used for long-term ANT-DBS were selected by demonstrating the activation of Papez circuit, as evidenced by presence of a ∼40 ms. latency HPC evoked response^79^ (see supplementary materials). Bipolar pairs were selected for chronic ANT-DBS to reduce stimulation artifacts and optimize HPC sensing for LPF seizure and IES detection (see supplementary data). Bipolar ANT electrode contact pairs were used for ANT-DBS with a continuous LF-DBS (2/7 Hz; 200us pulse width; 2 - 6 mA) or duty cycle (1 min on/ 5 min off) HF-DBS (145 Hz; 100 – 200 us pulse width; 2 - 5 mA). Participant M1, M3, M4, and M5 had periods of both LF-and HF-DBS. Participant M2 did not have adequate data rates during HF-DBS and was omitted from the General linear mixed model (GLMM) analysis (see supplementary materials).

### Long-term sensing local field potentials

Four bipolar AMG-HPC electrode pairs were selected for sensing based on ability to record seizures and IES. The four bipolar pairs can be selected from any of the 16 electrode contacts. The LFP sampling rate can be set at 250, 500, or 1000 Hz. Most of the LFP data was collected at 250 Hz because of significant increase in wireless data drops with higher 500 Hz and 1000 Hz sampling rates. (see supplementary materials)

### Local Field Potential Analysis

The LFP analysis was performed using the EPA and cloud platform with AI computational infrastructure and visualization. Validated automated machine-learning algorithms were applied to the long-term LFP recordings to identify seizures, IES^34^ and classify wake-sleep state (Awake, REM, and NREM)^39,40,50^. The algorithm pipeline identifies seizures, IES, brain impedance^80^, and sleep-wake behavioral state for consecutive 30-second data segments. The algorithms and their performances were previously reported^34,39^ (see supplementary materials). The sleep-wake classifications and seizure events are synchronized with patient reports for further offline analysis. Toolboxes and data for automated LFP analysis for detecting IES, seizures, and performing sleep-wake classification are freely available (https://github.com/bnelair/best_toolbox).

### Seizure and IES detection

The IES and seizures are electrographic biomarkers of pathologic, epileptogenic brain tissue and are readily identified by human visual review of LFP recordings. We used a previously validated algorithm^34,49^ for detecting IES transients from LFP recordings. The adaptive IES algorithm enables detecting IES in long term LFP data with changing background activity commonly encountered in prolonged recordings spanning week to months of time. Here the gold-standard training data was used to set a hypersensitive threshold for all participants (see supplementary materials).

The intracranial LFP associated with epileptic seizures exhibits characteristic temporal and spectral evolution over a wide frequency range. We previously described an accurate seizure detector developed using a convolutional neural network with long-short-term-memory (CNN-LSTM) utilizing the Short Time Fourier Transform of the LFP as the input^34^. The CNN-LSTM outputs a seizure probability for 10 second data segments. Gold-standard visually reviewed seizures are used for training, validation, and pseudo-prospective testing. The seizure detection performance using the CNN-LSTM model was previously reported for participants M1-4 and here is applied to all five participants using a hypersensitive threshold to ensure all seizures are detected for the analysis. We visually reviewed all candidate seizure events detected by the hypersensitive automated CNN-LSTM detector. The temporal distribution of seizures was determined by plotting the circular histogram of seizure onset times for all verified seizures across all participants.

### Sleep-wake classification from continuous LFP recordings

Sleep-wake classification in ambulatory participants living in their natural home environment was performed using an individualized behavioral state classifier. As described previously in humans and canines simultaneous LFP and gold standard sleep annotations (expert review by EKS) from polysomnography (PSG) were used to train the fully automated sleep-wake classifier^39,40^. A validated Naïve Bayes classifier using features extracted from the LFP was used for long-term tracking of sleep-wake behavioral state classification.

### Statistical analysis

We employed a generalized linear mixed model (GLMM) to examine the impacts of different therapy periods (Baseline, LF-DBS, HF-DBS) and behavioral states (awake/sleep) on monthly self-reported seizure rates, monthly confirmed electrographic seizure and hourly IES rates. The analysis was done in R^81^. See the details on data used and fitted models in the supplementary materials.

### Verbal Memory Assessment in Ambulatory subjects in natural home environment

Verbal memory was tracked in ambulatory participants using an established verbal memory paradigm^51,52,82^. The task was programmed on the epilepsy patient assistant (EPA) application (Figure 6A) and remotely performed with the participant at different times of the day during the week, at their preference. The ANT-DBS was turned off during the task. Investigators remotely activated the task on a mobile device, recorded and documented the verbal responses. In this task, participants were presented with a list of twelve proper nouns with the goal of later recalling them. Participants were instructed to commit these individual words to memory as they appeared one by one on a mobile device screen. Each word was displayed for a duration of 1600 ms., followed by a randomly varying blank interval of 750– 1000 ms. between words.

Immediately after the presentation of the final word in each list (during the encoding phase), participants engaged in a 20 second distractor task consisting of solving arithmetic problems. Following the completion of the distractor task, participants were tasked with verbally recalling as many of the twelve words as they could from the previously presented list, in any order, within a 30-second time frame (recall phase). Each session encompassed 25 sets of this encoding-distractor-recall procedure. The stimulation was reactivated upon successful completion of the task. Tasks performed after recent seizures were discarded.

### Immediate Mood Assessments

Mood assessments were integrated into the Epilepsy Patient Assistant (EPA) application, which was programmed to randomly query participants for their responses. Participants were prompted on a randomly selected day and time (10AM-6PM; one-to-four times a week) to complete the Immediate Mood Scaler 12 (IMS)^55^. The IMS is an ecological momentary assessment of 12 questions (7-point Likert scale) evaluating anxiety and depression symptoms in the moment. (see supplementary materials)

## Data Availability

The MRI, electrophysiology, and participant outcome data are available upon reasonable requests. A subset of awake, sleep, interictal and ictal continuous LFP recordings are available at the BNEL website.

## Code/Software/Data availability

All data and analysis code are available on reasonable request from the authors (https://github.com/bnelair/best_toolbox). MATLAB and Python code for the analysis and plotting figures for the main manuscript and supplementary information is available at https://github.com/bnelair

The IMS was presented on the EPA mobile device application through a licensing agreement with Posit Science (San Francisco, CA).

## Acknowledgements

This work was supported by National Institutes of Health–National Institute of Neurological Disorders and Stroke Grants UH2&3 NS095495, R01 NS112144 and U24 NS113637; Defense Advanced Research Projects Agency Grant HR0011-20-2-0028. The implanted devices were donated by Medtronic as part of the National Institutes of Health Brain Initiative. V.K. and V.S. were partially supported by institutional support of CTU in Prague. We thank Cindy Nelson, and Starr Guzman from Mayo Clinic for patient coordination; Abbey Becker, Dave Linde, and Scott Stanslaski from Medtronic for providing engineering support for the RC+S^TM^ devices and OpenMind for providing community expertise and resources (https://openmind-consortium.github.io/).

