## Supplementary data for "A platform for brain network sensing and stimulation with quantitative behavioral tracking: Application to limbic circuit epilepsy" (from another version of this preprint: version 2, 2024-02-28)

**Supplementary Data (text, tables, and figures)**

*Human participants.* Seven people were consented with drug resistant bilateral mesial temporal lobe epilepsy and five people (sTable 1) were implanted with an investigational neural sensing and stimulation device under FDA IDE: G180224 and Mayo Clinic IRB: 18-005483 *Human Safety and Feasibility Study of Neurophysiologically Based Brain State Tracking and Modulation in Focal Epilepsy*. The patients provided written consent in accordance with IRB and FDA requirements. Study registration https://clinicaltrials.gov/ct2/show/NCT03946618.

**Supplementary Table 1 (sTable 1) here**

**Protocol & Experimental Design**. Participant inclusion required demonstration of drug resistant mesial temporal lobe epilepsy (mTLE). For inclusion the patients must have had bilateral independent left and right temporal lobe onset seizures, or seizures from the dominant temporal lobe. Participants were required to have three or more disabling seizures per month as demonstrated on a mobile epilepsy patient assistant application (EPA) diary.

**Supplementary Figure 1 (sFig. 1) here**

Patients were required to have disabling focal aware seizures (FAS), focal impaired awareness seizures (FIAS) or focal to bilateral tonic-clonic seizures (FBTC) (Fisher et. Al. 2017). Seven patients were consented and five successfully completed the seizure diary baseline and were implanted with the investigational Medtronic Summit RC + S™ (RC+S) device (sFig. 2 & 3). Patients remained on stable medication regimes over the course of the study, except for M3 who was transitioned off a high-dose oral diazepam regimen due to sedation and fatigue and participant M5 who after implant was discovered to have frequent electrographic clinically necessitating increasing his Lacosamide and ultimately starting valproic acid.

**Primary Endpoints:**

1. Evidence that safety of EBS of ANT & HA is not worse than DBS in ANT and RNS in hippocampus for epilepsy as reported in SANTE Trial (Fisher R. et al. 2010) and NeuroPace Trial (Morrell M. et al. 2011).
2. Demonstration of 24/7-continuous iEEG monitoring and telemetry from the RC+S to the EPA, and feasibility of automated seizure detection and behavioral state classification using patient specific algorithms.
3. Demonstration of feasibility of tracking mood, anxiety and cognition using EPA device.

**Secondary Endpoints:**

1. Demonstrate accurate, automated seizure diaries.
2. Demonstrate characteristics of brain state dependent (wake, sleep, interictal, and pre-ictal, post-ictal) electrical stimulation evoked responses.
3. Quantification of iEEG biomarkers.
4. Quantification of iEEG biomarkers associated with behavioral state, mood, anxiety, and memory.
5. Demonstrate feasibility of brain state dependent (wake, sleep, interictal, and pre-ictal, post-ictal) adaptive electrical stimulation. We trialed continuous low frequency and high frequency ANT-DBS

**Ethics:** Prior to conducting the research, all protocols were approved by the Mayo Clinic Institutional Review Board (IRB). An IRB-approved study investigator or coordinator obtained informed consent from each participant (please see attached Informed Consent Form draft). Our institution uses an electronic submission process, and the IRB meets up to three times weekly ensuring a timely review / revision / approval process.

**Inclusion Criteria of Epilepsy Patients:** Adults with drug resistant bilateral mesial temporal lobe OR unilateral (dominant) mesial temporal lobe epilepsy were candidates. In these patients resective surgery has significant risk of memory impairment. All patients had bilateral medial temporal lobe epilepsy, or unilateral (dominant) temporal lobe epilepsy based on clinical evaluation using clinical semiology, scalp electroencephalography (EEG), and MRI neuroimaging. Patients that were not adequately localized with non-invasive studies had intracranial electroencephalography (iEEG) to demonstrate seizures are of mesial temporal origin.

1) Focal epilepsy, including seizures with and without impairment of consciousness, and secondarily generalized seizures:

i. disabling seizure counts > 3 per month on average over 1 – 3 months (Disabling seizures are those with significant negative impact on the patient's life)
ii. drug resistance to >2 appropriate seizure drugs with therapeutic serum concentrations
iii. not a good candidate for resective surgery or at significant risk for verbal memory decline as determined by our institution's multidisciplinary Epilepsy Surgery Committee.
iv. For 1 months prior to enrollment, participant's anti-seizure medication dosages have been stable, and participant has had at least 3 disabling (as defined in Inclusion Criteria 1) seizures per month, on average, with a seizure-free interval not to exceed 30 days. Seizures must be separated by a minimum of eight hours not to be considered part of a cluster. A cluster of seizures, for the purpose of this criterion, shall be considered a single seizure.

2) With the exception of epilepsy, participant had to be medically and neurologically stable.
3) Mayo Clinic Epilepsy Surgery Committee approval for brain stimulation therapy obtained on clinical grounds and without reference to this protocol.
4). Age 18 to 75.
5). Ability and willingness to provide informed consent and participate in the study protocol. Participant was able interpret and to respond, in accordance with the study protocol, to the advisory indicators provided by the device.
6) Participant had seizures that are distinct, stereotypical events that can be reliably counted by the patient or caregiver.
7) Participant could reasonably be expected to maintain a seizure diary alone or with the assistance of a competent individual.
8) Participant was able to complete regular office visits and telephone appointments in accordance with the study protocol requirements.
9) A female participant had to have a negative serum pregnancy test prior to the actual surgical procedure being done. If sexually active, must be using a reliable form of birth control, be surgically sterile, or be at least two years post-menopausal.
10) Participant's seizure focus, based upon clinical semiology, scalp encephalography (EEG) and neuroimaging, shall support bilateral temporal lobe epilepsy or unilateral (dominant) temporal lobe epilepsy. Patients that were not adequately localized with non-invasive studies had intracranial electroencephalographic (iEEG) to demonstrate seizures are of mesial temporal origin.
11) Participant were informed of his or her eligibility for resective surgery as a potential

alternative to the study, if such surgery is a reasonable option.
12) Participant speaks and reads English.
13) Participant have had a brain magnetic resonance imaging (MRI) epilepsy evaluation within the past two years.
14) Participant had iEEG documentation of ictal events consistent with his or her predominant current seizure type.
15) Participant's anatomy permitted implantation of the Medtronic Investigational RC+S generator within 20 mm of the skin surface.
16) Participant was expected to periodically check battery levels and recharge devices (Implanted neural stimulator (INS), Clinical Telemetry Module (CTM), and EPA mobile device) alone or with the assistance of a competent individual.

**Exclusion Criteria**: Patients was not enrolled if any of the following criteria apply:
1) For 3 months prior to enrollment, participant's anti-seizure medication dosages have not been stable, or participant has had more than 25 disabling (as defined in Inclusion Criteria 1) seizures per month, on average, or there was a seizure-free interval longer than 30 days within the past 3 months. Clinical seizures must be separated by a minimum of eight hours to not be considered part of a cluster. Cluster seizures are considered a single seizure event.
2) Participant has a contraindication to magnetic resonance imaging.
3) Participant has a substance abuse history (alcohol, prescription, or illicit medications) within the preceding two years.
4) Participant participated in another drug or device trial within the preceding 30 days.
5) Participant has been hospitalized for a psychiatric condition within the preceding two years or has had a history of psychosis within the preceding two years (excluding post-ictal psychosis).
6) Participant is implanted with pacemaker, implantable cardiac defibrillator, cardiac management product, or a medical device that interferes with the RC+S device. This includes, but is not limited to, direct brain neurostimulators, spinal cord stimulators, vagus nerve stimulators (VNS), and cochlear implants. Patients with a vagus nerve stimulator implanted but turned off through the duration of the study may be enrolled, provided their clinical status has been stable for at least one month with VNS turned off. Alternatively, patients with a VNS may have the previously disabled VNS removed at time of surgery to implant the Medtronic RC+S.
7) Participant has been diagnosed with psychogenic or non-epileptic seizures.
8) Participant has been diagnosed with primary generalized seizures.
9) Participant has experienced unprovoked status epilepticus in the preceding year.
10) Participant has had therapeutic surgery to treat epilepsy that may interfere with electrode placement.
11) Participant is on anticoagulants and is unable to discontinue them peri-surgically, as required by the neurosurgeon or Investigator.
12) Participant has significant platelet dysfunction from medical conditions or medications (including, particularly, aspirin or sodium valproate). If platelet dysfunction is suspected, participant can be enrolled only if a hematologist, the Investigator, and the neurosurgeon judge it to be advisable.
13) Participant is ineligible for cranial surgery.
14) Pregnancy

**Evaluations prior to enrollment**

All patients had: 1) Epilepsy monitoring unit evaluation to record the patient's habitual seizures with scalp video-EEG (31 to 76 scalp electrodes 2) 7T structural and functional MRI (unless 7T MRI is contra indicated, then 3T or 1.5T). The fMRI is to determine language lateralization. 3) Neuropsychological evaluation. 4) Presentation at a multidisciplinary epilepsy surgery conference 5) Based on the results of the pre-surgical evaluation and multidisciplinary epilepsy surgery conference patients may proceed to Wada study to lateralize language and access risk of memory impairment with surgery. Furthermore, patients may undergo invasive phase II monitoring with intracranial electrodes and prolonged video intracranial EEG monitoring for precise localization of their seizures if necessary. Patients with bilateral mesial temporal lobe seizures or dominant mesial temporal lobe seizures were candidates for this study.

**Evaluations repeated during study (sFig. 1)**
Participants were recording seizures and medications daily. In addition, mood, and anxiety and verbal [1] and spatial memory [2] tasks on EPA were collected when requested. The patient was also queried by the EPA to fill out electronic versions of the PHQ-9[3] and GAD-7[4] on their EPA as needed. The EPA data was transferred to the secure Mayo server and reviewed daily by the health care team.

Disabling seizures can be simple focal with and without impairment of consciousness, and secondarily generalized tonic-clonic seizures.  Investigators classified seizure types per the International League Against Epilepsy (ILAE) classification[5]. Seizure classification were initially conducted upon enrollment and then amended throughout the participant's participation in the study if new seizure types occurred.

In addition to EPA acquired data on seizures, medications, mood & anxiety, we utilized standardized outcome measures obtained in periodic face to face clinical visits. Standardized outcome measures[6] were obtained at beginning and periodically throughout study including: 1) Mood Assessments 2) Quality of life Assessments 3) Seizure severity assessments 4) Neuropsychological assessments.

**Seizure Event Diaries/Seizure Event Registration on EPA:** Patient-maintained seizure diaries are an accepted outcome measure in epilepsy-related therapeutic trials, including those evaluating brain stimulation[7,8]. Throughout the study, the epilepsy participants were asked to maintain a daily seizure journal on the EPA detailing the number, type, and estimated duration of seizures experienced each day. Participants were provided with and trained to use a commercially available smart watch and wearable devices.

**Home Medication Administration Record on EPA:** Participants used their mobile device to mark times of medication administration between study visits. Ongoing records of medication regimens were kept through the duration of the study.

**Assessments on the EPA that were collected as needed:**

1) Assessments of Mood & Anxiety obtained on the EPA include Immediate mood scaler, Generalized Anxiety Disorder 7 (GAD-7), Patient Health Questionnaire 9 (PHQ-9)

2) Assessments of Verbal & spatial memory on the EPA: Performance of verbal (FR, CatFR, PAL) & spatial (navigation) memory were tested at home by patient using the EPA on a volunteer basis.

Comprehensive outcome assessments were obtained at baseline, 3-6 months post-implant and 9-15 months post-implant. The instruments collected include the following:

1) Mood Assessments collected during office visits. Change in mood assessed by

- Beck Depression Inventory II – a 21-item self-reported instrument for measuring the presence and severity of depression in adults and adolescents. Each of the 21 items requires the respondent to endorse one of 4 options, scored 0 to 3, with increasing scores reflecting greater severity of a given depressive symptom.
- Columbia Suicide Severity Rating Scale (C-SSRS): a multiple question instrument administered by a health care professional for measuring suicidal ideation and suicidal behavior. The C-SSRS has 13 items with either yes-no responses or 5-point severity scores. The questions are organized progressively, and severity of suicidal ideation and behaviors are only explored if found to be present.

2) Quality of Life Assessments during office visits. Change in health-related quality of life (QOL) will be assessed using the following:

- Quality of Life in Epilepsy-31 (QOLIE-31) – a self-reported survey containing multi-item measures of quality of life, emotional well-being, role limitations, social support/isolation, energy/fatigue, worry about seizure, medication effects, health discouragement, work/driving/social function, attention/concentration, language, memory, physical function, pain, and health perceptions.
- SF-36 Health Survey (SF-36) – a widely used measure of health-related quality of life. The SF-36 contains 36 items that are divided into 8 different categories designed to evaluate the multidimensional aspects of health including: physical functioning, role limitations caused by physical problems, bodily pain, general health perceptions, vitality, social functioning, role limitations caused by emotional problems, and mental health. The 8 subscales can be aggregated into 2 summary scores, physical component summary and mental component summary, with higher scores representing better health status.

3) Seizure Severity Assessment during office visits. Seizure severity will be assessed using the following:

- Liverpool Seizure Severity Scale (LSSS) – a widely used measure of seizure severity (Cramer JA and French J, 2001). The Liver- pool Seizure Severity Scale is used to quantify the patient's perceptions of changes in seizure severity during the course of a clinical trial. The scale assesses the seizure severity based on two factors: patients' perceptions of control over their seizures and the perceived severity of the ictal and postictal phenomena. The scale consists of 16 items: percept of control subscale (percept), six items; perceived ictal/postictal severity (ictal/postictal), ten items. Each item is scored on a l-4-point Likert response scale.
- Seizure Severity Questionnaire (SSQ) Version 3 – a review of various aspects of seizures completed by the person who has seizures, possibly with help from people who have observed the seizures. There are 11 questions asking about events before, during, and after typical seizures. Participants who are seizure free following the device implantation procedure are not required to complete the SSQ.
- Mayo Seizure Scale – 5 questions answered by patient or caregiver to quantify seizure frequency.

4) Neuropsychological Assessments during office visits will include:

- Boston Naming Test – a 60-item confrontation naming test to measure word retrieval performance. The participant is asked to name what they see in a picture book, and the time it takes for them to respond is recorded.
- Controlled Oral Word Association Test (COWA) (FAS) – a measure of verbal fluency. Participants are given one minute to name as many words as possible beginning with the letter F. The procedure is then repeated for letters A and S.

- Rey Auditory Verbal Learning Test: is a test that evaluates verbal learning and memory (recall and recognition). Participants are given a list of 15 unrelated words repeated over five different trials and asked to repeat. Another list of 15 unrelated words are given and repeated. The participant must then repeat the original list of words and then again after 30 minutes.
- Wechsler Memory Scale (WMS-IV) Visual Reproduction Test – a measure of visual memory. Participants are shown an image for 10 seconds and then asked to draw the image.
- Wechsler Memory Scale (WMS-IV) Verbal Paired Associates – a measure of verbal memory for associated word pairs. The participant is read 10- or 14-word pairs. The participant is then read half the word pair and asked to respond with the corresponding other half of the pair.

Verbal memory was assessed using the Rey Auditory Verbal Learning Test (RAVLT)[9] and Wechsler Memory Scale – 4$^{th}$ Edition (WMS-IV) Verbal Paired Associates Subtest[10]. The WWMS-IV Visual Reproduction Subtest[10] was used to assess visual learning and memory. Language testing included the Boston Naming Test (BNT)[11] (Kaplan 2001) and the controlled oral word association test (COWAT, Letters CFL)[12]. The WMS-IV tests were scored utilizing the published test manual. To assess performance on the RAVLT, the Mayo Normative Studies published norms were utilized [13]. In instances where participants fell below the normative age range, the lowest age available was used. Performance on the verbal fluency and naming tests are presented as raw scores. Change in performance over time was defined as greater than 1.5 standard deviations in either direction compared to baseline.

**Supplementary Table 2. (sTable 2.) here.**

**BrainRISE Platform (sFig 2).** The electrophysiology data, patient reports and ambulatory verbal memory testing in participants in their natural home environments were collected using the epilepsy patient assistant (EPA), a custom software application running on a mobile device enabling bidirectional communication between implanted devices with wireless connectivity and local and cloud computing resources. The BrainRISE system enables synchronized[14] dense behavioral tracking and LFP brain activity to be collected (sTable 3). The EPA features include automated algorithms for continuous analysis of LFP data, control of electrical stimulation and impedance testing, LFP analysis and an interface for collecting patient interactions[15,16].

**Supplementary Figure 2. (sFig 2) and Table 3 (sTable 3) here**

**The Medtronic RC+S$^{TM}$ Summit Device (Supplementary sFig. 3).** The local field potential (LFP) data were acquired using the RC+S$^{TM}$ Summit. Each patient had four implanted leads targeting left and right anterior nucleus of thalamus (ANT), amygdala (AMG) and hippocampus (HPC). (sTable 4). The neural sensing-stimulation device enables programmable16 channel electrical stimulation, monopolar and bipolar 2-point impedance measurements, and continuous 4 channel (selected bipolar pairs) wireless streaming of brain local field potentials (LFP) to a mobile device and cloud environment[15]. The integration and synchronization of the RC+S$^{TM}$ Summit with off-the-body computing devices (mobile device, smart phone, watch) uses a custom software application (Epilepsy Patient Assistant: EPA)[16]. The RC+S$^{TM}$ requires ~1.5 hours of charging for continuous 24-hr electrical brain stimulation (EBS), and LFP and impedance timeseries wireless streaming to the mobile device. The telemetry antenna requires charging every 72 hours. The mobile device running the EPA application is maintained with wall power and has ~8 hours rechargeable battery. The on-device analytics and electrical stimulation do not require connectivity with the system. The ANT-DBS therapy remains active as long as the implanted RC+S$^{TM}$ device is charged. With continuous streaming of 4-channel LFP at 250 Hz and ANT-DBS the device required daily charging. When the RC+S$^{TM}$ reaches 30% battery status the LFP streaming is automatically disabled to maintain therapy. In this scenario with LFP streaming disabled therapy remain active for approximately 2 weeks.

**Supplementary Figure 3 (sFig.3) and Table 4 (sTable 4) here**

**Surgical Implantation:** Patients placed under anesthesia, a Leksell (Elekta) frame placed, and stereotactic MR images obtained. These data were then used to identify the anterior commissure (AC)/posterior commissure (PC) with COMPASS (COMPASS International) deep brain stimulation (DBS)–targeting software. Using a Schaltenbrand and Wahren atlas overlay and anatomical guidance[17] we targeted the dorsal medial and anterior nucleus[18]; however, we used a modified target based on anatomy to allow contact with dorsal medial thalamus and the anterior nucleus.

Medtronic 3387 electrodes (Used in SANTE Trial[7] were implanted in the thalamus and Medtronic 3391 electrodes were implanted into the long axis of the medial temporal lobe. We used bilateral hippocampal trajectories implants designed to span 2.5 cm of the hippocampus and 1 cm of the amygdala. Intraoperative fluoroscopy was used to ensure their accurate placement. After placement of the leads, the patient was taken immediately (while under anesthesia) for post-placement CT to confirm localization (sTable 4). After the electrode locations were confirmed, the patient was taken back to the operating theater. The leads then tunneled to the Medtronic RC+S device.

A single RC+S device was implanted in the left subclavicular region performed independently, and 60-cm pain lead (37087-60; Medtronic) extensions were used to enable channel isolation for recordings greater than that of the standard DBS extension. The leads from the right side of the head will traverse over the skull and then down the left side of the neck in parallel with the left side lead implants.

After the implant the patient was taken to the intensive care unit to recover and dismissed on Postoperative Day 1 to 2. During the operation and time in the intensive care the sensing capabilities of the device was interrogated, and the data recorded.

**MRI and Diffusion Tensor Imaging (DTI): Lead and Electrode Contact Localization**

All participants had a seizure protocol 3 Tesla MRI and DTI. Co-registration with post-op CT imaging was used to localize the electrodes within the ANT and AMG-HPC. Using frame stereotaxis bilateral ANT (Medtronic 3387 leads) and HPC & AMG (Medtronic 3391 leads) were implanted in 4 participants. Participant M2 was implanted with a 3387 rather than a 3391 in her residual left HPC from a prior anterior temporal lobectomy. The 3387 lead was used for its smaller contact spacing given the small posterior HPC tail remnant. The 16 electrode contacts were localized with post-operative CT co-registered to the pre-op MRI for anatomic localization using previously described pipelines[19,20]. The CT and electrode contact positions are co-registered to a $T_1$ weighted anatomical MRI scan using co-registration in SPM12 (https://fil.ion.ucl.ac.uk/spm/) and Freesurfer (http://surfer.nmr.mgh.harvard.edu/) was used to segment the $T_1$ MRI and the electrodes labeled according to the Destrieux atlas[21–23]. The final electrode contact localization for impedance analysis was performed with Lead DBS[20] (sFig. 3, sTable 4).

**ANT-DBS**

During the study continuous LF-DBS (2 & 7 Hz; 200us pulse width) and duty cycle HF-DBS (145 Hz; 100 & 200 us pulse width; 1 minute on-5 minute off) were investigated. For analysis we aggregate 2 & 7 Hz and 145 Hz. Subgroup analysis did not identify difference between the two LF-DBS and HF-DBS paradigm.

**Sensing Local Field Potentials (LFP)**

The LFP signals were continuously recorded while trialing two different stimulation paradigms applied to the anterior nucleus of the thalamus (ANT-DBS). The LFP signals were recorded with sampling frequency of 250 Hz or 500 Hz. The sampling frequency was determined as a compromise between data quality, wireless data transmission dropout, and device battery limitations (sTable 3).

**Hippocampal Evoked potentials**

To identify ANT-DBS stimulation contacts that engage the limbic circuitry we used ANT-DBS with concurrent hippocampal (HPC) sensing as previously described to identify ANT stimulation electrodes[24].

**Supplementary Figure 4 (sFig. 4) here**

**Automated LFP Analysis of Epileptiform Activity and Behavioral State**
We deployed a previously developed suite of automated tools for sleep classification[25], interictal epileptiform spike (IES) and seizure detection[14] synchronized with patient reports of seizure, mood and sleep using the EPA[15,16]. The system is integrated with a cloud computing environment providing visualization of continuous LFP data, brain state classification, IES rates, seizures, and the information entered by patients. The automated electrographic seizure detection algorithm running on the mobile device and in the cloud identifies electrographic seizures using high sensitivity thresholds. The candidate seizure events are subsequently visually reviewed and confirmed or rejected by a human expert.

**Behavioral state classification**
We used a previously validated algorithm developed from simultaneous polysomnography (PSG) and LFP recordings[25]. Expert visual review was used to create gold standard labeled training, validation and testing data as defined by AASM: Awake, Rapid-eye-movement (REM) sleep, and three non-REM (N1-N3) sleep stages. The automated classifier using LFP signals recorded from AMG, ANT, and HPC produced accurate classification (average F1-score 0.89 on testing data) and was subsequently deployed in the five patients for behavioral state classification (Awake, REM, Non-REM (N2 & N3). (sTable 5 & 6).

**Supplementary Figure 5 (sFig. 5) and sTable 5 & 6 here**

**Statistical analysis of seizures and IES**
Candidate seizures were identified using a hypersensitive detector applied to the LFP recordings as previously described[14]. The quantification of epileptiform activity (seizures and

IES) are shown on polar histograms for left and right mTLE seizures. We investigated whether low-frequency or high-frequency ANT-DBS paradigms differentially effected the occurrence of left and right mTLE seizures. There was no difference detected (sFig. 6 & 7).

The hippocampal LFP data were analyzed using a system previously described for tracking electrographic seizures and interictal spikes[14](sTable 3, 7 & 8). All electrographic events were detected by a highly sensitive automated detector applied to the hippocampal LFP data, followed by manual elimination of false positive detections[14]. We employed a generalized linear mixed model (GLMM) to examine the impacts of different therapy periods (LF-DBS, HF-DBS) and behavioral states (awake/sleep) on monthly self-reported seizure rates, monthly confirmed IEEG seizure rates, and hourly IES rates.

**Supplementary Figure 6 (sFig. 6) and Table 7 (sTable 7) here**

Electrographic events exceeding 10 seconds in duration and exhibiting ictal electrophysiological markers (such as low voltage fast activity, periodic spiking with a frequency greater than 2 Hz, and evolution in amplitude or frequency, followed by postictal silencing) were classified as seizures. We documented and confirmed electrographic seizures in all 5 subjects. In several cases, seizures involved both HPCs in each patient (sTable 8). These confirmed seizures were not subclassified based on any time series analytical methods. Despite variability in seizure onset, all seizures were treated uniformly in subsequent statistical analyses. Seizures occurring within an hour of each other were grouped as a single clustered seizure in 4 patients. Patient M4 experienced multiple seizures during 3-day periods, interspersed with seizure-free weeks. For M4, seizures were grouped in 72-hour intervals to reflect this unique clustering pattern. We calculated circadian seizure distribution from these clustered seizures for each patient. To analyze the impact of behavioral states, seizures were categorized based on the time of occurrence; those during average sleep hours were assigned to the sleep state, and others to the awake state. Clustered seizures were tallied over 4-week periods, with weekly steps and 3-week overlaps. The GLMM evaluated these monthly (4-week) seizure rates, with and without consideration of behavioral states, to assess the impact of sleep and ANT-DBS therapy on electrographic seizures.

**Supplementary Table 8 (sTable 8) here**

We utilized an automated spike detector[26], validated on a portion of the current dataset[14], across all HPC channels. The hourly spike rates for each patient were calculated for all HPC channels and then adjusted based on the data transmitted during that hour. Rates were divided by the data rate, excluding hourly segments with less than 50% data rate and segments where a seizure occurred within the hour. Night and day segments with less than 3 hours of data were also omitted. Sleep and awake spike rates were estimated as medians from intervals between 12 am - 8 am and 9 am - 10 pm, respectively. The final dataset comprised estimated hourly spike rates in different behavioral states (sleep and awake), adjusted for transmitted data.

Patients used the epilepsy patient application (EPA) to log seizures as they occurred, or retrospectively if logging was forgotten or not possible at the time. Self-reported seizures were collected during the pre-implant period (6-12 weeks) and continuously throughout the study, covering periods of stimulation OFF and open-label ANT-DBS therapy. These seizures were clustered using the same intervals as for iEEG seizures (72-hour for M4; 1-hour for M1, M2, M3, and M5). Clustered seizures were counted in 4-week intervals with a 3-week overlap, considering only seizures from a specific therapy period and discarding overlapping intervals between consecutive therapy periods. If the final interval within a therapy period was 3 weeks, seizure counts were normalized to a 4-week interval.

**Statistical analysis methods**

We analyzed hourly spike rate data and monthly seizure rate data using a generalized linear mixed model (GLMM) via the lmer package in R. Models for self-reported seizures, iEEG seizures, and IES were fitted using the glmer.nb function, which employs a negative binomial distribution for data fitting. The models were fitted using maximum likelihood (Laplace Approximation) and the Nelder-Mead optimizer. Fitted models provided estimates and standard errors for fixed effect variables, with z-value significances calculated from these estimates and standard errors. We full model (with random intercepts and slopes). Each model type was analyzed twice, with and without 7 Hz LF-DBS data (sTable 9, sTable 10). We investigated the effect of omitting M3 data since M3 did not undergo HF-DBS therapy (refer to supplementary table 11 for model comparison).

The self-reported seizure model included study periods (Baseline, NOSTIM, LF-DBS, HF-DBS) as the sole categorical predictor, with the number of self-reported seizures in 4 weeks as the dependent variable. The subjects were the grouping variable of GLMM. The model included all four study periods as fixed and random effects.

The EEG seizure model assessed the average 4-week seizure rate during LF-DBS and HF-DBS therapy periods, with these periods serving as categorical predictors. The dependent variable was the number of confirmed iEEG seizures in 4 weeks. The full model used ANT-DBS (LF/HF) periods as random effect variables, further extended with different data fitted to estimate seizure rates during sleep or awake, seizures with left or right onset, and unilateral or bilateral seizures (sFig. 7), under different therapies (LF-DBS, HF-DBS). See the results in sTable 11.

**Supplementary Tables 9, 10 & 11 here.**

The spike rate model estimated group-level IES rates during sleep and awake states, and across different therapy periods (LF-DBS/HF-DBS). The hourly IES rate was the dependent variable. Fitted data points were daily median IES rate during sleep and awake hours. Both models included three fixed effect variables: ANT-DBS (LF/HF), behavioral states (sleep/awake), and EEG channel laterality (left/right). The model employed a nested grouping structure, with the main group being the subjects, each having a nested group of all their iEEG channels. The model's random effect variables were the behavioral states (sleep/awake) and therapy periods (LF-DBS, HF-DBS).

**Supplementary Figure 7 (sFig. 7)**

**Memory Assessment with free-recall task Mood Assessments in Ambulatory Participants**

**Free recall task:** We used a Free Recall Task to assess participants' verbal memory[27]. The participants completed the tasks on a mobile device in their home environments while iEEG and behavioral data were streamed to the cloud repository. Each participant with lists of 12 randomly chosen nouns, displayed individually. After a distractor task of basic addition, the participant was asked to recall as many of the words as possible. Each session consisted of 15 novel lists. ANT-DBS was paused during the task period. A researcher remained on the phone with the participant for the duration of the task.

**Ecological momentary assessments:** One-to-four times a week, participants were prompted on a random day and time to complete the Immediate Mood Scaler 12 (IMS)[28]. The IMS is a 12-item ecological momentary assessment with 7-point Likert scale questions evaluating severity of anxiety and depression symptoms in the moment. Higher total IMS scores indicate less severe symptoms, and lower (more negative) scores indicate more severe symptoms. The IMS was presented through the mobile device through a licensing agreement with Posit Science (San Francisco, California). The IMS scores were averaged within each week for comparison with the single, weekly, memory score.

**Statistical analysis:** For group-wise, statistical comparisons, we used a two-sample Wilcoxon Rank Sum test (wilcox.test in R; R Core Team R. R: A language and environment for statistical computing. 2013). In group-wise comparisons, free recall verbal memory scores were significantly higher during periods of low frequency stimulation than during high frequency stimulation ($p < 0.001$). Of note, stimulation was paused for the duration of the task, so participants were not actively receiving stimulation at that time. The average weekly IMS score did not differ significantly between high and low frequency stimulation ($p > 0.05$). Although these results are suggestive of an impact of stimulation frequency on memory, further evaluation is required to determine if possible confounding factors such as a difference in seizure frequency at different stimulation parameters might account for this effect. The absence of pre-stimulation, baseline IMS and Free-Recall results because the BrainRISE system was not available at the time of enrollment.

### Supplementary Tables

#### sTable 1. Patient Demographics and Clinical Information.

| Participant | Age, Hand(L/R), Gender (M/F), Onset age & DRE (yrs.) | Risk Factor (RF) Medical Hx: Surgical Hx: | TL Seizure Types | ASM 1) Current 2) Previous | EEG 1) Scalp 2) Intracranial | MRI PET SPECT |
|---|---|---|---|---|---|---|
| **M1** | 55-60 y.o.; RF.<br>Onset 5-10. y.o.<br>DRE: 36 yrs.<br>EMU: scalp EEG & sEEG. | RF: TBI<br>Medical: Anxiety & Depression<br>Surgical: None | FAS, FIAS, Rare FBTC | 1) GBP, CZP<br>2) LEV, OXC PHT, CBZ | 1) B.IES & Szs.<br>2) B.HPC IES & Szs | MRI: Normal<br>PET: N/A<br>SPECT: N/A |
| **M2** | 20-25 y.o.; RF.<br>Onset 5-10 y.o.<br>DRE 3 yrs.<br>EMU: scalp EEG only | RF: None<br>Medical: Anxiety, Depression, DM.<br>Surgical: L.ATL | FAS, FIAS, Rare FBTC | 1) LCM, CZP,<br>2) LEV, PHT, CBZ, OXC | 1) B. IES & Sz.<br>2) N/A | MRI: L.ATL<br>PET: N/A<br>SPECT: N/A |
| **M3** | 40-45 y.o.; RF.<br>Onset 30-35 y.o.<br>DRE 10 yrs.<br>EMU: scalp EEG & iEEG | RF: None<br>Medical: Anxiety, Depression, GAD-65 positive serum/CSF<br>Surgical: VNS | FAS, FIAS, Rare FBTC | 1) CZP, LEV, CNB<br>2) LZP, CBZ, PHT, TPX, LCM, CBD | 1) B.IES & Szs.<br>2) B.HPC IES & Szs | MRI: Normal<br>PET: N/A<br>SPECT: N/A |
| **M4** | 35-40 y.o.; RF.<br>Onset 0-5 y.o.<br>DRE 7 yrs.<br>EMU: scalp EEG & sEEG | RF: None<br>Medical: Anxiety, Depression, GAD-65 positive serum/CSF<br>Surgical: None | FAS, FIAS, Rare FBTC | 1) OXC, LEV<br>2) LCM, CBZ, PHT, LGT | 1) B.IES & Szs.<br>2) B. HPC IES & Szs | MRI: Normal<br>PET: N/A<br>SPECT: N/A |
| **M5** | 30-35 y.o. RM.<br>Onset 20-25 y.o.<br>DRE 5 yrs.<br>EMU: scalp EEG only | RF: TBI & Family history of epilepsy<br>Medical: Anxiety, Depression<br>Surgical: None | FAS, FIAS, Rare FBTC | 1) LCM, VPA<br>2) LEV, LGT, PHT, CBZ | 1) B.IES & Szs.<br>2) N/A | MRI: L. HPC atrophy & incr. T2 signal<br>PET: N/A<br>SPECT: N/A |

**Supplementary Table 1. Patient Demographics:** The patients had mesial temporal lobe epilepsy and HPC seizures. All patients had bilateral independent IES and seizures in the Phase-1 evaluation and M1, M3 and M4 had invasive stereo-EEG (sEEG) evaluations and had bilateral independent IES and seizures.

Abbreviations: Electroencephalography (EEG). Drug resistant epilepsy (DRE). Male/Female (M/F). Right/Left (R/L). Bilateral (B). Deep Brain Electrical Stimulation (DBS). Traumatic brain injury (TBI). Vagus Nerve Stimulation (VNS). Anterior temporal lobectomy (ATL), Temporal lobe (TL), Focal aware seizures (FAS), Focal impaired awareness seizures (FIAS). Focal to bilateral tonic-clonic (FBTC). Gabapentin (GBP), Cenobamate (CNB), Clonazepam (CZP), Levetiracetam (LEV), Lorazapam (LZP), Oxcarbazepine (OXC), Phenytoin (PHT), Carbamazepine (CBZ), Lacosamide (LCM), Lamotrigine (LGT), Valproic Acid (VPA). Interictal epileptiform spikes (IES), Seizures (Sz), increased (incr.), Hippocampus (HPC).

**sTable 2. Participant Device Related Adverse Events & Outcomes.**

| | | AE | QOLiE | BDI | LSSS | BNT | CFL | RAVLT | RAVLT (DR) | VPA (IR) | VPA (DR) | VR (IR) | VR (DR) |
|---|---|---|---|---|---|---|---|---|---|---|---|---|---|
| M1 | Baseline | 0 | 50.1 | 10 | 65 | 59 | 37 | 47 | 26 | 5 | 7 | 9 | 6 |
| | Low Frequency | 0 | 34.8 | 26 | 58 | 52 | 32 | 36 | 19 | 4 | 4 | 7 | 6 |
| | High Frequency | 1 | 30.1 | 18 | 68 | 56 | 35 | 36 | 15 | 4 | 4 | 7 | 5 |
| M2 | Baseline | 0 | 49.2 | 27 | 48 | 41 | 36 | 31 | 23 | 7 | 5 | 8 | 5 |
| | Low Frequency | 1 | 42.7 | 38 | 49 | 40 | 32 | 35 | 27 | 5 | 5 | 9 | 9 |
| | High Frequency | NA | NA | NA | NA | NA | NA | NA | NA | NA | NA | NA | NA |
| M3 | Baseline | 0 | 41.3 | 18 | 55 | 50 | 32 | 43 | 44 | 11 | 9 | 6 | 8 |
| | Low Frequency | 0 | 44.5 | 21 | 60 | 48 | 25 | 44 | 32 | 5 | 4 | 7 | 7 |
| | High Frequency | 0 | 41.6 | 19 | 38 | 51 | 24 | 52 | 40 | 6 | 5 | 10 | 7 |
| M4 | Baseline | 0 | 46.1 | 16 | 80 | 51 | 45 | 21 | 18 | 8 | 4 | 10 | 6 |
| | Low Frequency | 0 | 66.7 | 3 | 58 | 53 | 39 | 13 | 18 | 8 | 5 | 10 | 6 |
| | High Frequency | 1 | 53.4 | 9 | 53 | 48 | 30 | 5 | 11 | 3 | 2 | 10 | 7 |
| M5 | Baseline | 1 | 60.6 | 2 | 45 | 56 | 36 | 47 | 44 | 11 | 9 | 10 | 9 |
| | Low Frequency | 0 | 53.5 | 8 | 43 | 58 | 37 | 52 | 44 | 10 | 11 | 9 | 10 |
| | High Frequency | 0 | 46.6 | 13 | 35 | 56 | 43 | 56 | 44 | 11 | 12 | 9 | 10 |

**Supplementary Table 2. Participant Device Related Adverse Events & Outcomes.** Participants were monitored during baseline and while receiving low-frequency (LF-DBS) or high-frequency (HF-DBS) brain stimulation (average 99.2 +- 43 weeks). There were 4 device related adverse events reported to the FDA. Participants M1 and M4 developed worsening dysphoria, anxiety and sleep disruption with continuous HF-DBS. The symptoms resolved after changing to duty cycle (1 minute on & 5 minute off) HF-DBS parameters. M2 inadvertently disabled therapy using the epilepsy patient assistant (EPA) application. A subsequent EPA reversion corrected this potential for operator error, and the cloud dashboard more clearly displayed the therapy status to identify clinical team. At surgery for M5 the device lead could not be completely seated into the connector leaving on 3 recording right amygdala hippocampus (HPC) contacts. There were no infections, intracranial hemorrhages, status epilepticus, or deaths. Participant M4 was hospitalized for new onset functional event. Participant M5 had 3 overnight hospitalizations with recurrent seizures like during the baseline. All participants had repeated neuropsychological testing (NP) during the trial. Testing took place during scheduled in-person office visits. All participants had NP at baseline and during LF-DBS and HF-DBS. There were no significant group level changes in NP assessments. However, for participants M3 and M5 there were decreases in VPA immediate and delayed recall compared to baseline and in RAVLT for M5 during HF-DBS.

Abbreviations: Anterior nucleus of the thalamus (ANT); Boston Naming Test (BNT; Raw Total of 60); Letter Fluency Raw (CFL); Deep brain stimulation (DBS); Hippocampus (HPC); Rey Auditory Verbal Learning Test (RAVLT); Verbal Paired Associates (VPA); Visual Reproduction (VR). Delayed Recall (DR); Immediate Recall (IR).

**sTable 3. BrainRISE Platform Performance.**

| Participant | Weeks Monitored | Daily LFP data (hr.) | Total Reported seizures | EPA Reported seizures/month |
|---|---|---|---|---|
| **M1** | 168 | 15.16 ± 3.03 | 144 | 3.43 ± 2.28 |
| **M2** | 92 | 5.64 ± 3.58 | 151 | 6.57 ± 4.51 |
| **M3** | 80 | 17.09 ± 2.11 | 429 | 21.45 ± 7.27 |
| **M4** | 104 | 16.97 ± 2.29 | 59 | 2.27 ± 0.78 |
| **M5** | 52 | 13.92 ± 3.22 | 30 | 2.31 ± 3.01 |
| ***Mean*** | *99.2±43.0* | *13.76 ± 4.73* | *162±158* | *7.2 ± 8.1* |

**Supplementary Table 3. BrainRISE Platform Performance.** Participants were monitored over multiple months while receiving anterior nucleus of the thalamus deep brain stimulation (average 99.2 +- 43 weeks). Daily wireless local field potential (LFP) data transmitted varied considerably across participants and reflects current technology limitation, and challenges with managing multiple rechargeable wireless streaming devices. Participants reported their seizures using the epilepsy patient assistant (EPA) application.

**sTable 4. Electrode Contacts Localization.**

| | Lead 1<br>Left ANT | | | | Lead 2<br>Left AMG-HPC | | | | Lead 3<br>Right ANT | | | | Lead 4<br>Right AMG-HPC | | | |
|---|---|---|---|---|---|---|---|---|---|---|---|---|---|---|---|---|
| | $E_0$ | $E_1$ | $E_2$ | $E_3$ | $E_4$ | $E_5$ | $E_6$ | $E_7$ | $E_8$ | $E_9$ | $E_{10}$ | $E_{11}$ | $E_{12}$ | $E_{13}$ | $E_{14}$ | $E_{15}$ |
| **M1** | THL | THL | THL | THL | PHPC | HPC | HPC | HPC | THL | THL | THL | THL | AMG | HPC | HPC | HPC |
| **M2** | THL | THL | THL | THL | PHPC | PHPC | PHPC | HPC | THL | THL | THL | THL | AMG | HPC | HPC | HPC |
| **M3** | - | THL | THL | THL | AMG | HPC | HPC | HPC | THL | THL | THL | THL | AMG | AMG | HPC | HPC |
| **M4** | THL | THL | THL | THL | AMG | HPC | HPC | PHPC | THL | THL | THL | THL | AMG | HPC | HPC | HPC |
| **M5** | THL | THL | THL | THL | - | HPC | HPC | HPC | THL | THL | THL | THL | HPC | HPC | HPC | - |

**Supplementary sTable 4. Electrode Contact Localization**. Multiple limbic network nodes were targeted. Lead DBS localization of bilateral Amygdala and Hippocampus (AMG & HPC), Anterior Nucleus Thalamus (ANT) leads and electrode contacts. Localization of 16 electrode contacts for participants M1-5. The four leads were stereotactically targeted to left THL (contacts: $E_0$, $E_1$ $E_2$ $E_3$), left AMG-HPC (contacts: $E_4$, $E_5$ $E_6$ $E_7$), right THL (contacts: $E_8$, $E_9$ $E_{10}$ $E_{11}$), and right AMG-HPC (contacts: $E_{12}$, $E_{13}$ $E_{14}$ $E_{15}$). Table adapted from Mivalt et al. 2023[29].

**sTable 5. Sleep-wake classification scoring accuracy.**

| Participant | ANT-DBS | Awake accuracy [%] | NREM accuracy [%] | REM accuracy [%] | Total accuracy [%] |
|---|---|---|---|---|---|
| **M1** | | | | | |
| | No | 98.3 | 93.5 | 93.4 | 92.6 |
| | LF | 99.5 | 94.1 | 94.6 | 94.1 |
| | HF | 95.8 | 99.3 | 95.6 | 95.3 |
| **M2** | | | | | |
| | No | 95.1 | 96.3 | 95.5 | 93.5 |
| | LF | 95.2 | 96.3 | 95.4 | 93.5 |
| **M3** | | | | | |
| | No | 93.1 | 94.4 | 94.4 | 90.9 |
| | LF | 92.8 | 93.4 | 94.6 | 90.4 |
| | HF | 95.0 | 94.5 | 94.2 | 91.9 |
| **M4** | | | | | |
| | No | 89.0 | 91.3 | 93.5 | 86.9 |
| | LF | 84.1 | 90.3 | 92.9 | 83.7 |
| | HF | 86.8 | 91.6 | 90.6 | 84.5 |
| **M5** | | | | | |
| | No | 94.7 | 92.1 | 93.7 | 90.3 |
| | LF | 93.5 | 94.4 | 89.1 | 88.6 |
| | HF | 97.9 | 98.8 | 96.8 | 96.8 |
| **Overall** | No | 94.0 ± 3.0 | 93.5 ± 1.8 | 94.1 ± 0.8 | 90.8 ± 2.3 |

**Supplementary Table 5. Automated classification of sleep and wake across different parameters of stimulation.** The average classification accuracy of Awake, NREM, and REM was 90.8% across all participants.

**sTable 6. Sleep characteristics.**

| Sleep | M1 | M2 | M3 | M4 | M5 | Overall |
|---|---|---|---|---|---|---|
| **Onset** [hr:min] | 12:27 am ± 01:07 | 02:18 am ± 02:22 | 10:43 am ± 00:39 | 10:42 pm ± 00:37 | 01:07 am ± 02:07 | 11:59 pm ± 01:56 |
| **Offset** [hr:min] | 08:55 am ± 01:01 | 11:33 am ± 03:01 | 07:39 am ± 00:31 | 08:06 am ± 00:51 | 08:29 am ± 02:59 | 08:35 am ± -2:07 |
| **Sleep Duration** [hr:min] | 08:19 ± 01:28 | 08:49 ± 03:04 | 08:49 ± 00:40 | 09:28 ± 00:52 | 07:07 ± 02:01 | 08:33 ± 01:56 |
| **NREM Duration** [hr:min] | 05:39 ± 00:57 | 07:01 ± 02:21 | 06:29 ± 00:37 | 06:16 ± 00:54 | 03:06 ± 00:45 | 05:41 ± 01:51 |
| **REM Duration** [hr:min] | 01:58 ± 00:29 | 01:09 ± 00:40 | 02:09 ± 00:27 | 02:41 ± 00:52 | 02:35 ± 01:06 | 02:10 ± 00:56 |
| **Awake Duration** [hr:mins] | 00:42 ± 00:40 | 00:39 ± 00:33 | 00:11 ± 00:12 | 00:30 ± 00:39 | 01:26 ± 01:05 | 00:41 ± 00:48 |
| **Cycles** | 5.13 ± 2.43 | 2.78 ± 2.47 | 3.90 ± 2.55 | 3.67 ± 2.07 | 5.00 ± 2.00 | 4.04 ± 2.50 |
| **NREM Epoch Duration** [mins] | 18.46 ± 29.36 | 28.16 ± 40.59 | 20.53± 33.04 | 16.89 ± 8.71 | 23.20 ± 27.02 | 19.93 ± 31.91 |

**Supplementary Table 6. Average Sleep Characteristics.** The participant specific sleep-wake classifier was trained in the epilepsy monitoring unit using gold-standard scalp polysomnography and streaming local field potentials from hippocampus (see supplementary materials). The average sleep onset times, offset times, sleep duration, total non-rapid eye movement (NREM) duration, total rapid eye movement (REM) duration, total awake duration, number of NREM-REM cycles, and average duration of NREM epochs were tabulated for all participants. The average sleep duration for all 5 participants was 08:33 ± (01:56) with 4:04 ± 2.50 NREM-REM cycles. The average NREM epoch duration was 19.93 ± 31.91 minutes. The table highlights the variability across and within individual participants.

**sTable 7. Reviewed seizures vs all detected events.**

| Participant | Detected Events | Reviewed Seizures | Seizures/Detected Events [%] |
|---|---|---|---|
| **M1** | 1769 | 821 | 46.4 |
| **M2** | 7745 | 18 | 0.2 |
| **M3** | 564 | 440 | 78.0 |
| **M4** | 869 | 45 | 5.2 |
| **M5** | 8519 | 228 | 2.0 |
| **Total** | 19466 | 1552 | |
| **Mean** | 3893±3904 | 310 ± 331 | 26 ±34 |

**Supplementary Table 7. Reviewed seizures vs detected events.** Number of automated seizure detections in prospectively deployed hypersensitive detector in participants living with epilepsy in their home environments compared to manually reviewed seizures.

**sTable 8. Lateralization of Epileptiform Activity.**

| **Lateralization of interictal epileptiform spikes (IES) and seizures** | | | | |
|---|---|---|---|---|
| Participant | IES L>R (**p<0.001 all participants**) | | Seizure L>R (**p<0.001 M1,2,4,5**) | |
| | L. AMG-HPC | R. AMG-HPC | L. AMG-HPC | R. AMG-HPC |
| M1 | 1,928 ± 1,061 | 199 ± 276 | 690 | 75 |
| M2 | 1,830 ± 2,247 | 1603 ± 929 | 3,730 | 18 |
| M3 | 1,859 ± 859 | 969 ± 567 | 171 | 269 |
| M4 | 265 ± 246 | 37 ± 28 | 45 | 0 |
| M5 | 1,273 ± 482 | 725 ± 548 | 179 | 48 |

**Supplementary Table 8. Lateralization of Epileptiform Activity.** Comparison of interictal epileptiform spike (IES) rates and number of seizure onsets in post-implant ambulatory monitoring phase of the study. All participants had seizures that started unilaterally and then propagated to contralateral amygdala-hippocampus (AMG – HPC). All participants had bilateral independent IES, and all but M4 had bilateral independent seizures recorded. In all participants the IES activity was increased in left compared to right AMG-HPC. Similarly, M1, M2, M4 and M5 had more seizures originating from the left AMG-HPC. Note that M4 did not have seizures from right AMG-HPC during tracking in natural home environment and medication regime in contrast to the hospital epilepsy monitoring evaluation.

**sTable 9. Intracranial recorded Seizures used in GLMM.**

| Participant | ANT-DBS | Total seizures used in GLMM | Behavioral state | | Seizure onset | | | Seizure propagation | |
|---|---|---|---|---|---|---|---|---|---|
| | | | awake | sleep | Left | Right | Bilateral | Uni-lat. | Bi-lat. |
| M1 | LF | 304 | 284 | 20 | 239 | 34 | 31 | 172 | 132 |
| | HF | 117 | 102 | 15 | 114 | 1 | 2 | 64 | 53 |
| M2 | LF | 11 | 11 | 0 | >3730* | 11 | 0 | - | - |
| | HF | - | - | - | - | - | - | - | - |
| M3 | LF | 36 | 30 | 6 | 23 | 13 | 0 | 36 | 0 |
| | HF | 214 | 191 | 23 | 73 | 141 | 0 | 201 | 13 |
| M4 | LF | 11 | 11 | 0 | 11 | 0 | 0 | 11 | 0 |
| | HF | 15 | 15 | 0 | 15 | 0 | 0 | 10 | 5 |
| M5 | LF | 94 | 59 | 35 | 71 | 22 | 1 | 83 | 11 |
| | HF | 134 | 120 | 14 | 108 | 26 | 0 | 119 | 15 |
| Total | | 936 | 823 | 113 | 654 | 248 | 34 | 696 | 229 |

**Supplementary Table 9. Intracranial recorded seizures used in GLMM.** Model fit with data from different ANT-DBS stimulation frequency where stimulation amplitudes were >= 3.5 mA, and low-frequency ANT-DBS (LF - 2Hz) and high-frequency ANT-DBS (HF – 145Hz). Seizures are categorized based on behavioral state (sleep/awake) and left, right or bilateral hippocampus seizure onset. And based on propagation as unilateral (Uni-lat.) or bilateral (Bi-lat).

**sTable 10. Seizures used in GLMM.**

| Participant | ANT-DBS | Total seizures used in GLMM | Behavioral state | | Seizure onset | | | Seizure propagation | |
|---|---|---|---|---|---|---|---|---|---|
| | | | awake | sleep | Left | Right | Both | Uni-lat. | Bi-lat. |
| M1 | LF | 667 | 608 | 59 | 539 | 74 | 54 | 374 | 293 |
| | HF | 154 | 135 | 19 | 151 | 1 | 2 | 97 | 57 |
| M2 | LF | 18 | 18 | 0 | >3730* | 18 | - | - | - |
| | HF | - | - | - | - | - | - | - | - |
| M3 | LF | 51 | 42 | 9 | 28 | 23 | 0 | 51 | 0 |
| | HF | 389 | 340 | 49 | 143 | 246 | 0 | 372 | 17 |
| M4 | LF | 12 | 12 | 0 | 12 | 0 | 0 | 12 | 0 |
| | HF | 33 | 32 | 1 | 33 | 0 | 0 | 20 | 13 |
| M5 | LF | 94 | 59 | 35 | 71 | 22 | 1 | 83 | 11 |
| | HF | 134 | 120 | 14 | 108 | 26 | 0 | 119 | 15 |
| Total | | 1552 | 1366 | 186 | 1085 | 410 | 57 | 1128 | 406 |

**Supplementary Table 10. Seizures used in GLMM.** Model fit with data from different ANT-DBS stimulation frequency where stimulation amplitudes were >= 3.5 mA, and low-frequency ANT-DBS (LF - 2Hz and 7Hz) and high-frequency ANT-DBS (HF – 145Hz). Seizures are categorized based on behavioral state (sleep/awake) and left, right or bilateral hippocampus seizure onset. And based on propagation as unilateral (Uni-lat.) or bilateral (Bi-lat).

**sTable 11. GLMM results**

| Data used in GLMM | Total number of monthly intervals over LF and HF ANT-DBS | | Fitted GLMM p-values LF-DBS vs HF-DBS | |
|---|---|---|---|---|
| | All LF + HF | 3.5+ mA 2 Hz LF | All LF + HF | 3.5+ mA 2 Hz |
| Awake + Sleep | 364 | 229 | **0.03** | **0.03** |
| Awake + Sleep (with M2) * | 454 | 255 | 0.27 | 0.16 |
| Awake | 364 | 229 | **0.03** | **0.01** |
| Sleep | 364 | 229 | 0.49 | 0.69 |
| Left onset | 364 | 229 | 0.24 | 0.33 |
| Right onset | 264 | 175 | 0.70 | 0.90 |
| Unilateral | 364 | 229 | 0.41 | 0.31 |
| Bilateral | 364 | 229 | 0.07 | 0.07 |

**Supplementary Table 11. GLMM results.** Models fitted with awake or awake and sleep data (M1, M3, M4, and M5) had lower predicted monthly seizures at a group level in LF-DBS compared to HF-DBS ($p<0.05$). *Model used M3 LF-DBS data.

**Supplementary Figures**

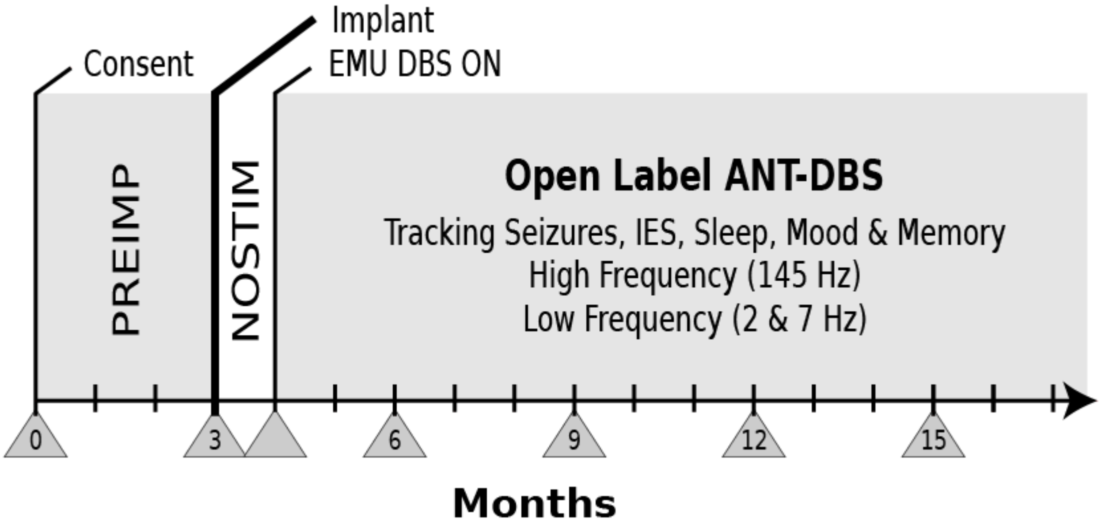


**Supplementary Figure 1. (sFig. 1). Safety & Feasibility Trial of Papez Circuit Tracking and Control.** The gray triangles denote in person office and hospital visits. Time 0 is the beginning of the study at time of consent for the study. Time 0 to 3 months is the diary phase where participants demonstrate use of EPA to catalog seizures and medications. Data on charging and maintaining devices is collected. At 3 months participant implanted with RC+S$^{TM}$ system. In the weeks following implant (~ 4 weeks post implant) patient admitted to epilepsy monitoring unit (EMU) for recording simultaneous polysomnography and intracranial EEG, i.e. local field potentials (LFPs), from hippocampus and anterior nucleus of thalamus. The participant leaves EMU with low frequency ANT-DBS. After 3-6 months of optimization of low ANT-DBS the participant transitioned to high frequency duty cycle ANT-DBS.  Neuropsychological data, quality of life, mood, and seizure severity collected at specified in person clinic follow-ups.

### Supplementary Figures

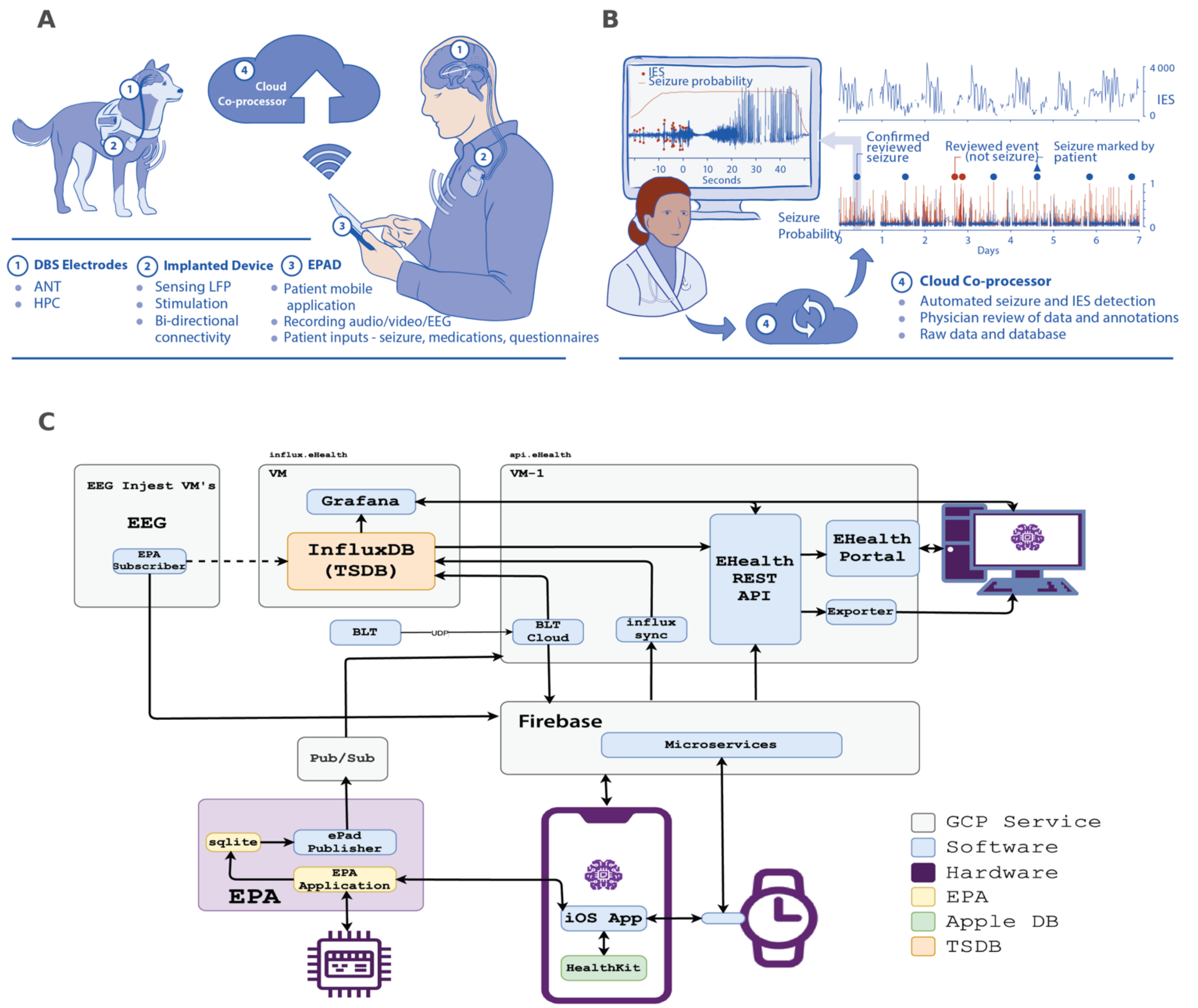


**Supplementary Figure 2 (sFig. 2). BrainRISE Platform for Optimizing Epilepsy Care.** Distributed brain co-processor integrating implanted sensing and stimulation devices with off-the-body and cloud computing resources. The system has been prospectively tested in canines and humans with drug-resistant epilepsy living in their natural environments. **A)** Schematic for bidirectional data transmission between implanted brain sensing and stimulation device integrated with a local mobile device running epilepsy patient assist (EPA) application and cloud computing environment. Deep brain-stimulation (DBS) electrodes implanted in critical network nodes of the participants epilepsy can be wirelessly addressed for stimulation and sensing. A particular use case of interest is limbic network epilepsy with electrodes targeting anterior nucleus of thalamus (ANT) and hippocampus (HPC). The BrainRISE platform provides chronic ambulatory monitoring with patient reported behavior, seizures, device data (battery, telemetry, and electrode impedance), and quantification of detected seizures, interictal epileptiform spikes (IESs) and wake-sleep behavioral states. Adapted from Sladky et. Al 2022[14]. **B)** The mobile device EPA application enables connection to multiple distributed devices and the cloud environment enables remote reviewing of electrophysiology data and analytics from a suite of algorithms running on the local mobile device or in the cloud environment. The physician can quickly review and confirm or reject automatically detected and patient reported candidate seizure events. The panel shows continuous hippocampal IES rates and seizure detection probability. Triangles show patient reported seizure events. Circles denote automated seizure detections either confirmed as seizures (blue dots) or false positive (red) by expert visual review. Monitor inset shows example of raw data from HPC with automated IES detections (red dots). The patient was aware and reported (triangle) one out of the six seizures detected in the continuous intracranial EEG (iEEG) and confirmed by the BrainRISE Dashboard. **C)** Schematic of integration of iPhone and iWatch with the platform running on Google Cloud running AI TensorFlow analytics, Firebase database, and Influx application for data viewing.

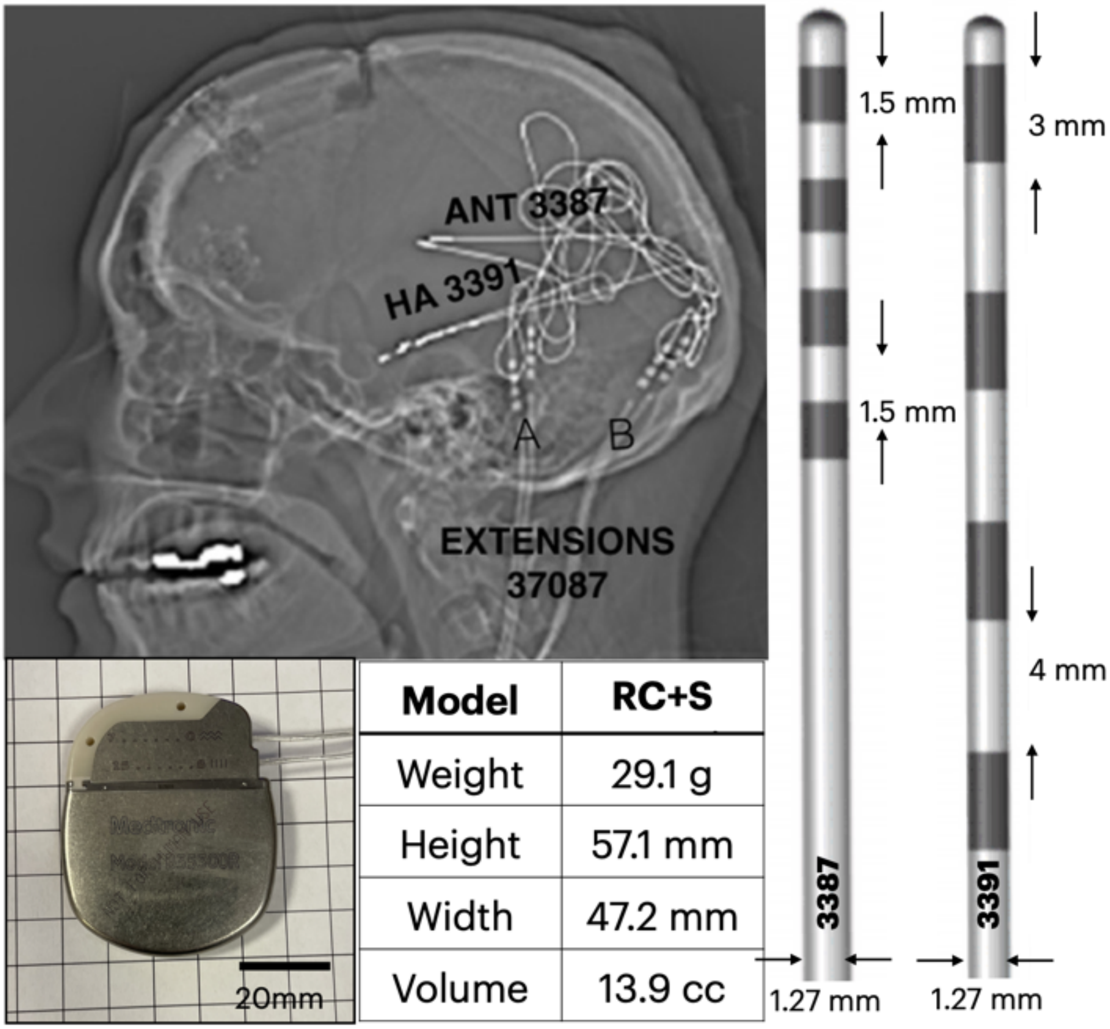


**Supplementary sFigure 3 (sFig. 3). Limbic Network Implant for Mesial Temporal Lobe Epilepsy (mTLE)**. The Medtronic RC+S$^{TM}$ enabled continuous tracking of limbic network electrophysiology and behavior. **A)** Lateral x-ray (M1) showing 3387 electrodes in the bilateral ANT and 3391 electrodes in bilateral AMG and HPC. **B)** The 3387 four contact (contact surface area = 5.985 mm$^2$) lead span 10.5 mm. The contacts are 1.5 mm long each and separated by 1.5 mm. The 3391 four contact (surface area = 11.97 mm$^2$) lead span 24.5 mm. The contacts are 3.0 mm long and separated by 4.0 mm. **C)** RC+S$^{TM}$ Summit implantable device dimensions. The rechargeable device enables 16 stimulation channels and programmable 4 sensing channels from any bipolar contact pair.

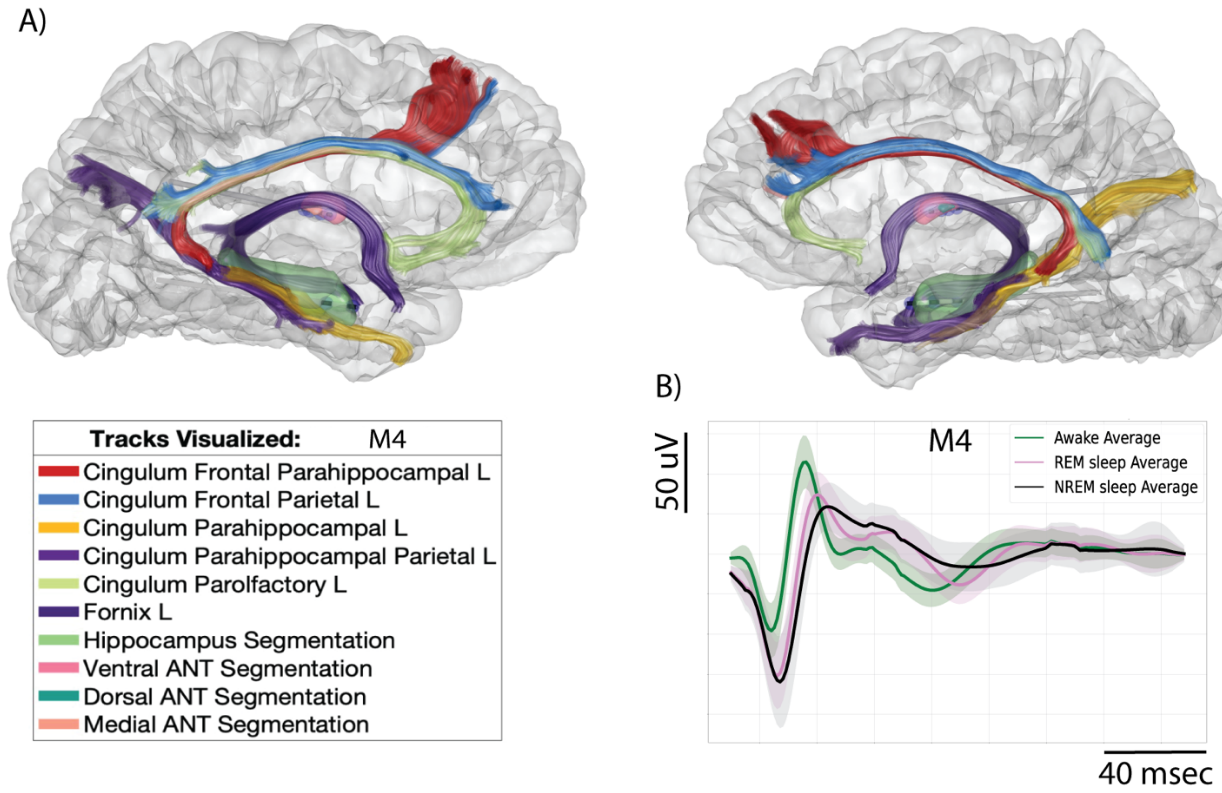


**Supplementary Figure 4 (sFig. 4). Electrode Contact Localization and DTI imaging of brain connectome**. Hippocampal (HPC) and anterior nucleus of thalamus (ANT) nodes were targeted here for patient M4. **A)** DBS localization of bilateral Amygdala (AMG) and Hippocampus, Anterior Nucleus Thalamus (ANT) leads and electrode contacts. Electrode Contact Localization to Pre-Op MRI and DTI imaging. **B)** Continuous 2 Hz stimulation of ANT produced evoked responses (ERP) sensed in HPC. Average HPC evoked potential response were used to identify electrode for stimulation. A HPC evoked response to ANT stimulation was used as a measure of network engagement here showing averaged responses during wake, non-rapid eye movement sleep (NREM), and rapid eye movement sleep (REM). Data are averaged across 24-hour segment showing average ERP (bold lines) and standard deviation (shaded areas) or ERP curves.

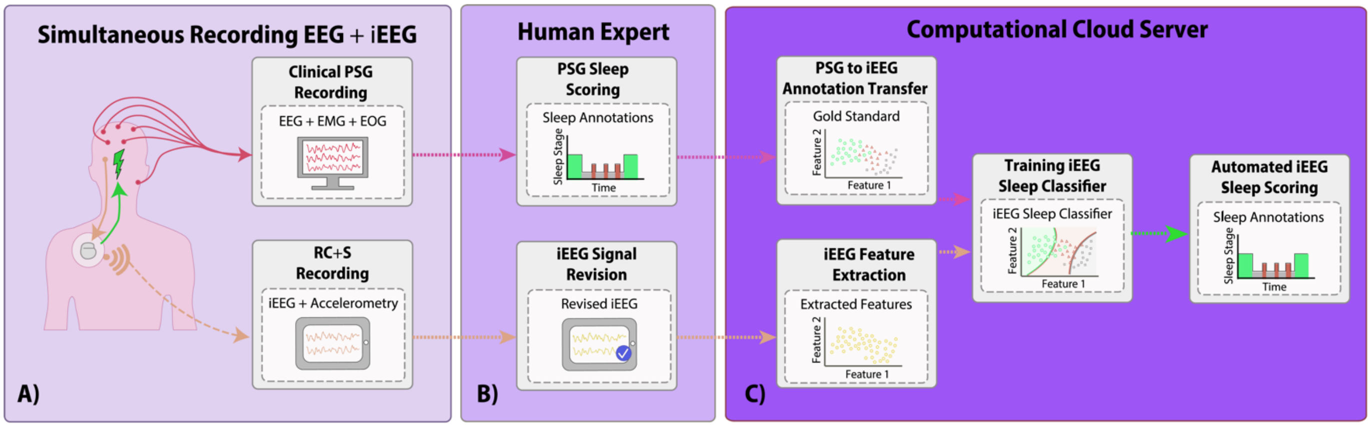


**Supplementary Figure 5 (sFig. 5). Naive Bayes Behavioral State Classification**
Block-diagram of the process for training, validation, and testing of an automated sleep classifier using simultaneous polysomnography (PSG) and intracranial recorded local field potentials (LFP) from hippocampus. **A)** Schematic of the simultaneous scalp and hippocampal LFP recordings. **B)** The expert gold-standard sleep scoring from PSG is used to create labeled data to develop the automated wake-sleep classifier. The hippocampal LFP recordings were reviewed and the channel with the lowest epileptiform activity and stimulation related artifacts was selected for use in the classifier **C)** Direct comparison of the expert sleep scoring are used to evaluate performance of the automated Naive Bayes LFP based sleep-wake classifier. Classifier is then deployed on long-term iEEG data only. The performance of the classifier is provided in supplementary sTable3. Illustration adapted from Mivalt 2022[25].

**Supplementary Figure 6**

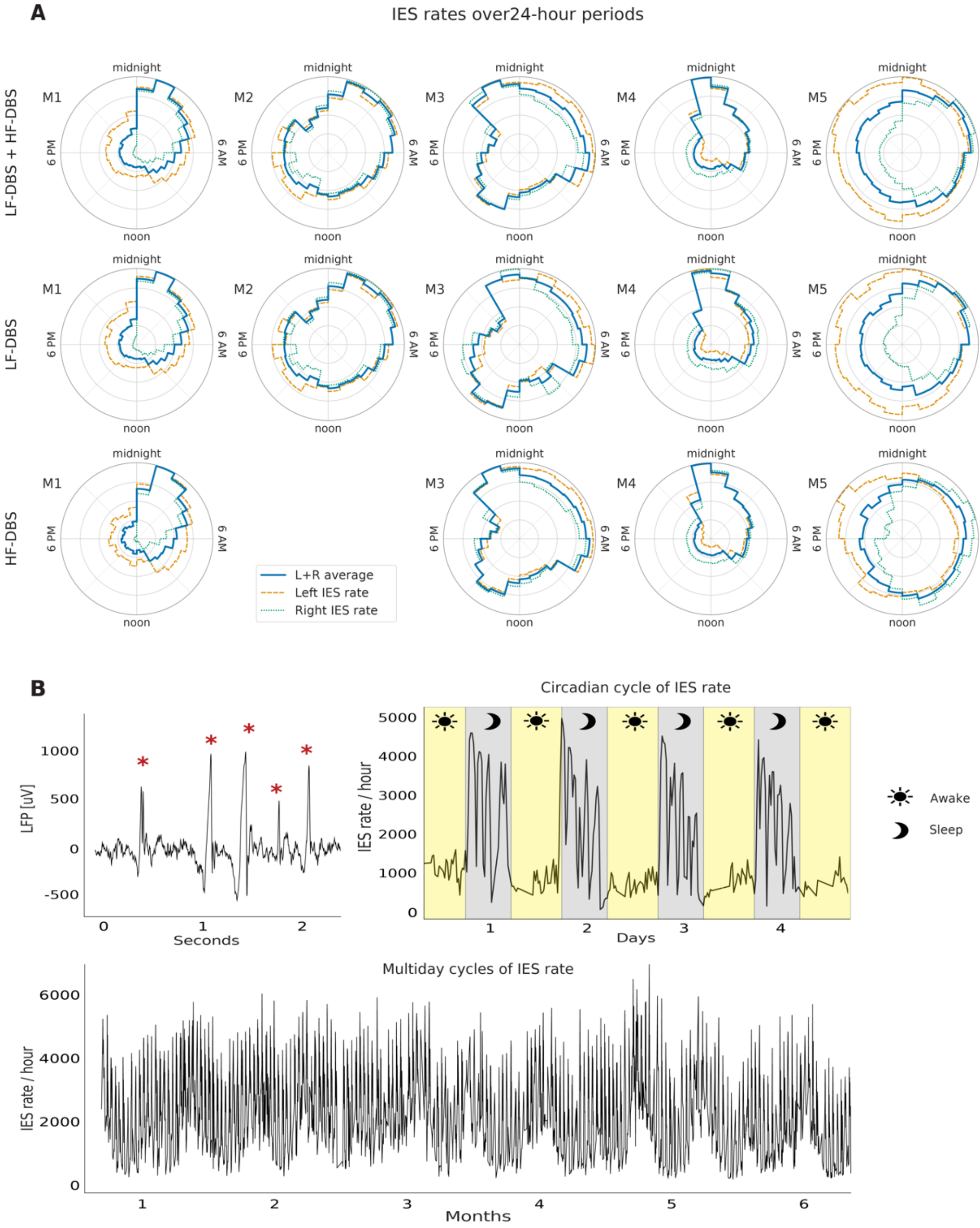


**Supplementary Figure 6 (sFig. 6) Interictal Epileptiform Spikes (IES) over 24-hour periods. A)** The IES rates are increased in sleep compared to wakefulness overall. There are interesting nuances however, with M9 showing greater or equal left temporal IES in wakefulness compared to sleep. The differences in IES distributions with different ANT-DBS paradigms is currently an area of investigation. **B)** M1 IES rates. The IES The pattern of IES show a crescendo – de-crescendo pattern after sleep onset at correlates with slow-wave (NREM2 & 3) sleep duration, which is greater during the early phases of sleep. The mechanism(s) of the behavioral state dependent excitability remains unclear, but may reflect synaptic[30] glia[31] or extracellular space changes[29].

**Supplementary Figure 7**

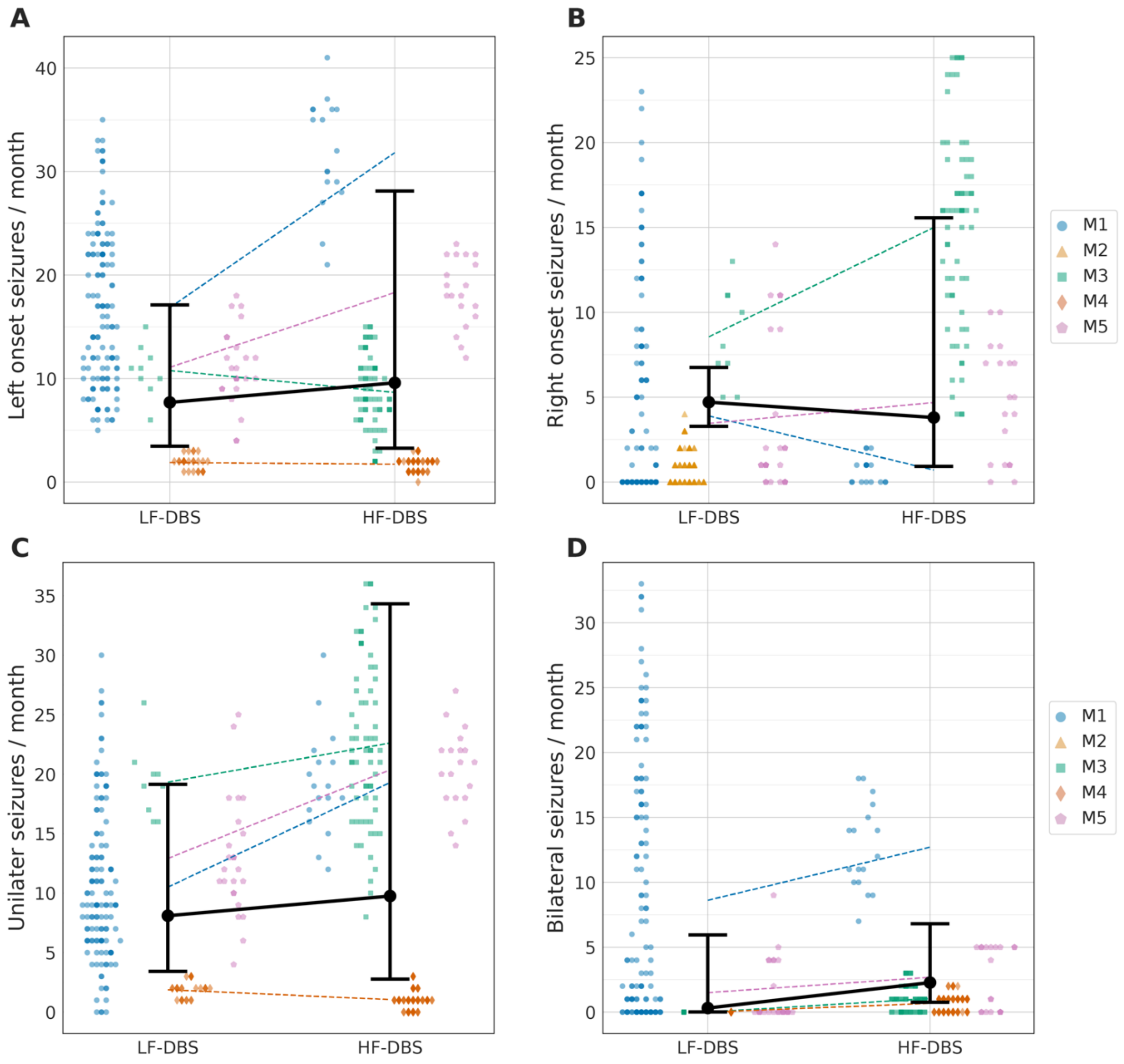


**Supplementary Figure 7 (sFig. 7). Left onset and right onset seizures during LF and HF ANT-DBS.** Sub analysis of individual left (A) and right (B) side didn't support a difference between LF and HF. Sub analysis of unilateral (C) and bilateral (D) seizures showed positive slopes between LF and HF but weren't significant. Data fit in the model are described in supplementary Table 6
